# Motor Skill Retention Impairments in Parkinson’s Disease: A Systematic Review with Meta-analysis

**DOI:** 10.1101/2022.12.18.22282724

**Authors:** Jacopo Cristini, Zohra Parwanta, Bernat De las Heras, Almudena Medina-Rincon, Caroline Paquette, Julien Doyon, Alain Dagher, Simon Steib, Marc Roig

## Abstract

The ability to acquire and retain motor skills is essential for persons with Parkinson’s Disease (PD), who usually experience a progressive loss of mobility during the disease. Deficits in the rate of motor skill acquisition have been previously reported in these patients. Whether motor skill retention is also impaired is currently not known. We conducted a review that included 46 studies to determine whether, compared with neurologically intact individuals, motor skill retention is impaired in PD. Meta-analyses revealed that, following a single practice session, persons with PD have deficits in skill retention (SMD = −0.17; 95% CI = −0.32, −0.02; *p* = 0.0225). However, these deficits are task-specific, affecting sensory motor (SMD = −0.31; 95% CI −0.47, −0.15; *p* = 0.0002) and visuomotor adaptation (SMD = − 1.55; 95% CI = −2.32, −0.79; *p* = 0.0001) tasks, but not sequential fine motor (SMD = 0.17; 95% CI = −0.05, 0.39; *p* = 0.1292) and gross motor tasks (SMD = 0.04; 95% CI = −0.25, 0.33; *p* = 0.7771). Importantly, retention deficits became non-significant when augmented feedback during practice was provided. Similarly, additional sessions of motor practice restored the deficits observed in sensory motor tasks. Meta-regression analyses confirmed that retention deficits were independent of performance during motor skill acquisition, as well as the duration and severity of the disease. These results are in line with prominent neurodegenerative models of PD progression and emphasize the importance of developing targeted interventions to enhance motor memory processes supporting the retention of motor skills in people with PD.

## 1. Introduction

Parkinson’s disease (PD) is one of the most common neurodegenerative disorders and the number of persons with this clinical condition is expected to double by 2040 (Dorsey et al., 2018). Parkinson’s is a complex, heterogeneous and progressive disorder, characterized by several motor and non-motor symptoms but its diagnosis is based on the onset of the cardinal motor features of the disease (Kalia and Lang, 2015). Pharmacological treatments are the first line of action to manage the motor symptoms of the disease but with time they tend to progressively lose efficacy (Connolly and Lang, 2014). As a result, patients with PD experience a relentless deterioration, leading to major motor dysfunctions, and eventually loss of autonomy (Aarsland et al., 2000). Implementing non-pharmacological interventions to maintain the functional independence of these patients is thus important (Keus et al., 2007).

Motor rehabilitation helps patients with PD maintain the motor skills needed to function independently (Abbruzzese et al., 2016). Motor learning, defined as the ability to learn, adapt, and retain long-term skilled movements (Kantak and Winstein, 2012) is the base of motor rehabilitation (Wolpert et al., 2011). Motor learning comprises skill acquisition and retention. Acquisition, the on-line process during which sensory and motor information is encoded through motor practice, is characterized by fast gains in skill performance during the initial phases of practice, followed by slower improvements (i.e., automatization) in later phases (Karni et al., 1998a). Skill retention, in contrast, is the result of an off-line process, during which the sensory and motor information is consolidated to form motor memories (Dayan and Cohen, 2011; Doyon et al., 2009a; Dudai, 2004). Skill retention is inferred through either change in skill performance measured during retention tests or by the ability to generalize the acquired skills to other tasks assessed with transfer tests. Retention is clinically important because it reflects the permanent ability of the patient to perform a motor skill and not transient improvements in skill performance (Kantak and Winstein, 2012; Schmidt et al., 2018).

Previous studies investigating deficits in skill retention in PD have reported inconsistent results (Marinelli et al., 2017a; Nieuwboer et al., 2009; Olson et al., 2019). Furthermore, the extent to which the task nature, amount of practice, type of feedback provided during motor practice, progression of the disease, and effect of antiparkinsonian medications moderate the capacity to retain motor skills has yet to be determined. This review, conducted following PRISMA guidelines (Moher et al., 2009) and registered in PROSPERO (<u>CRD42020222433</u>) (Booth et al., 2012), aimed to summarize the evidence regarding deficits in skill retention in people with PD relative to neurologically intact (NI) individuals. Determining to what extent people with PD have deficits in skill retention could stimulate the design of more individualized and effective motor rehabilitation therapies.

## 2. Methods

### 2.1. Eligibility criteria

The eligibility criteria of the studies included in the review were operationalized with the PECOS (population, exposure, comparator, outcomes, study design) framework (Morgan et al., 2016; Morgan et al., 2018). **Population**: participants with PD without other neurological comorbidities and not receiving deep brain stimulation (Marinelli et al., 2017a). **Exposure**: having idiopathic PD. **Comparator**: NI individuals of similar age. **Outcomes**: skill retention and transfer measured ≥ 1 h following the end of practice to capture long-term change (Dudai, 2004; Kantak and Winstein, 2012). **Study design**: observational studies with a PD and a NI group or interventional studies with a group of PD patients and NI individuals who did not receive the intervention.

### 2.2. Search strategy

Two authors performed independently the electronic search on electronic databases (Web of Science, PubMed, Scopus, PsycINFO, SPORT Discuss) and screened the reference lists of relevant reviews (Abbruzzese et al., 2009; Aslan et al., 2021; Barry et al., 2014; Clark et al., 2014; Felix et al., 2012; Hayes et al., 2015; Knowlton et al., 2017a; Krakauer et al., 4 2019; Marinelli et al., 2017a; Nackaerts et al., 2019; Nieuwboer et al., 2009; Olson et al., 2019; Ruitenberg et al., 2015; Siegert et al., 2006) as well as articles reviewed at the full-text level (see study selection section). The electronic search was neither language nor date restricted, but it was limited to peer-reviewed articles. The primary search was performed using the following three main terms and their variations: “Parkinson’s disease” (population), “healthy control” (comparator), and “motor learning” (outcome), combined with Boolean operators, and can be found in the supplementary (**Suppl. 1**). The final search was completed on December 2^nd^, 2021.

### 2.3. Study selection and data extraction

Two authors screened the list of titles and abstracts of articles retrieved in the search and selected potentially relevant articles for a more detailed review at full-text level. Following the screening of the articles, both authors held a meeting to compare their results. Disagreements were resolved by discussion including a third author.

Two authors extracted the following data from the studies: study design, number and characteristics of participants, characteristics of the motor task, as well as the outcomes and the endpoints used to assess motor learning. Means and standard deviations (SDs) of motor skill acquisition and retention/transfer test scores were extracted. Subsequently, both authors compared their data to confirm that they were entered correctly.

When an article did not provide means and SDs and this information could be inferred from figures, data were extracted using a web-based tool (https://WebPlotDigitizer/). When this was not possible, the authors of the study were contacted. If data could still not be obtained, the study was not included in the quantitative meta-analysis and results were reported qualitatively.

### 2.4. Methodological quality assessment

Risk of bias at the study level was assessed by two authors that used the NIH Quality Assessment Tool for Observational Cohort and Cross-Sectional Studies (National Heart and Institute, 2019). This tool, which comprises 14 items, has shown good validity to assess the methodological quality of observational studies (Ma et al., 2020). Given the design of the studies included in the review, items 10 (exposure repeatedly measured over time) and 12 (assessors blinded to the exposure) were scored as “not applicable” and not considered for evaluating study quality (National Heart and Institute, 2019) (**Suppl. 2**). The two authors rated each item as “yes”, “no”, or “not reported”. All responses other than “yes” indicate a risk of bias. The number of “yes” responses was used to calculate a percentage score (i.e., number of “yes”/12 * 100) and categorize studies as “good” (≥ 90%), “fair” (≥70, but <90), or “poor” (<70%) (National Heart and Institute, 2019). Sources of bias and heterogeneity were investigated with funnel plots and Egger’s regression tests (Egger et al., 1997; Page et al., 2019; Sterne et al., 2011).

### 2.5. Main analysis and influence of moderators

We grouped studies that assessed skill retention following a single session of practice or after extended practice (≥2 sessions) because retention tests following single and extended practice reflect different stages of the motor memory formation process (Dayan and Cohen, 2011). Subgroup meta-analyses investigated whether the nature of the task influenced skill retention. To this end, studies were classified following well-established motor learning classifications (Dayan and Cohen, 2011; Dijkstra et al., 2020; Doya, 2000; Doyon et al., 2009a; Hardwick et al., 2013; Hikosaka et al., 2002; Knowlton et al., 2017b; Krakauer et al., 2019; Maas et al., 2008; Masapollo et al., 2021; Segawa et al., 2015; Spampinato and Celnik, 2021; Surgent et al., 2019; Taylor and Ivry, 2012) as sensory motor (SMT), sequential fine motor (SQT), visuomotor adaptation (VAT), gross motor (GMT), and speech motor (SPT) tasks (**Table 1**). The influence of augmented feedback (Marinelli et al., 2017a; Schmidt et al., 2018) was assessed by grouping studies based on whether extrinsic feedback was provided or not, and if so, which type: knowledge of results, performance, or both (Schmidt et al., 2018).

**Table 1.**
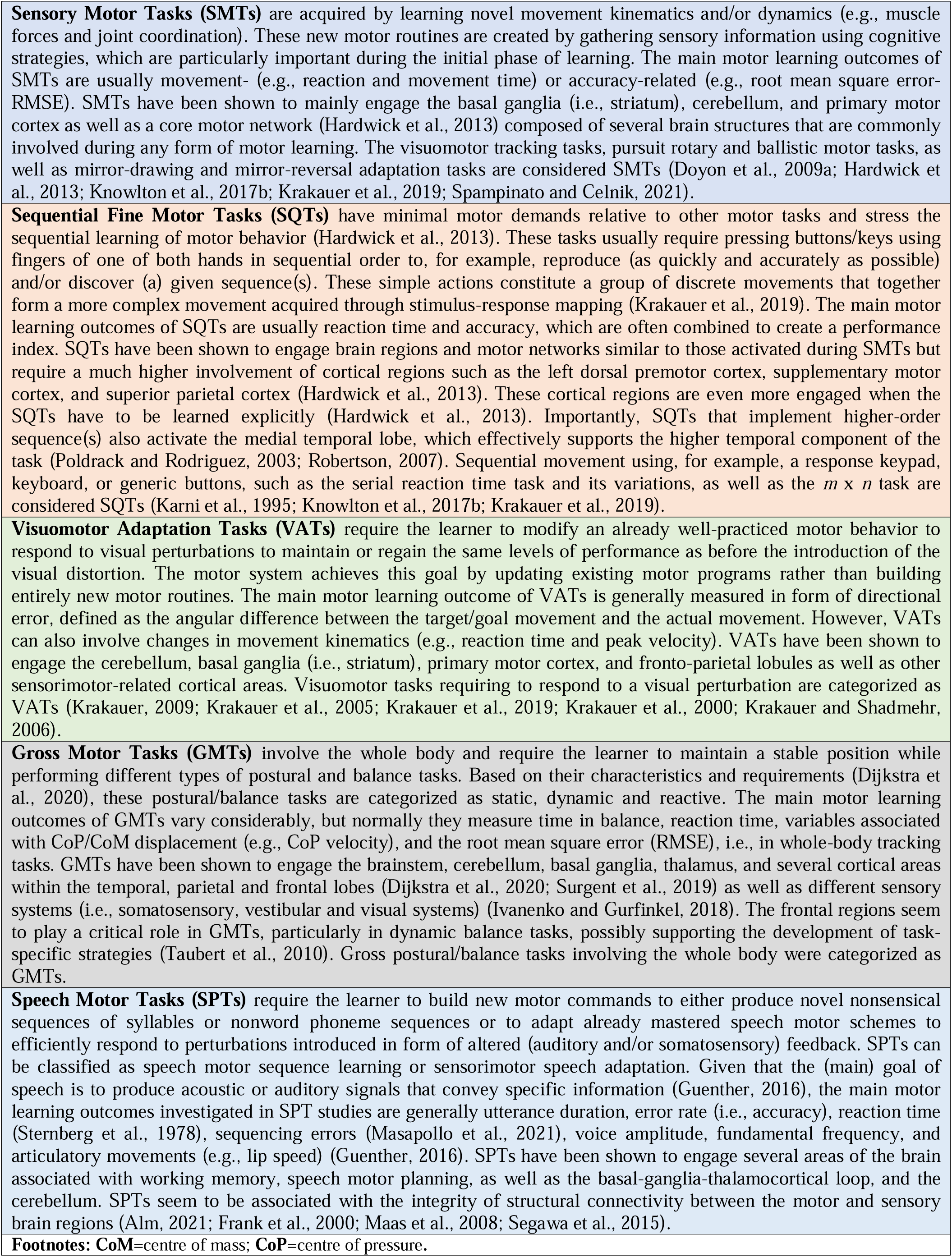
Task nature classification

|  |
| --- |
| **Sensory Motor Tasks (SMTs)** are acquired by learning novel movement kinematics and/or dynamics (e.g., muscle forces and joint coordination). These new motor routines are created by gathering sensory information using cognitive strategies, which are particularly important during the initial phase of learning. The main motor learning outcomes of SMTs are usually movement- (e.g., reaction and movement time) or accuracy-related (e.g., root mean square error-RMSE). SMTs have been shown to mainly engage the basal ganglia (i.e., striatum), cerebellum, and primary motor cortex as well as a core motor network (Hardwick et al., 2013) composed of several brain structures that are commonly involved during any form of motor learning. The visuomotor tracking tasks, pursuit rotary and ballistic motor tasks, as well as mirror-drawing and mirror-reversal adaptation tasks are considered SMTs (Doyon et al., 2009a; Hardwick et al., 2013; Knowlton et al., 2017b; Krakauer et al., 2019; Spampinato and Celnik, 2021). |
| **Sequential Fine Motor Tasks (SQTs)** have minimal motor demands relative to other motor tasks and stress the sequential learning of motor behavior (Hardwick et al., 2013). These tasks usually require pressing buttons/keys using fingers of one of both hands in sequential order to, for example, reproduce (as quickly and accurately as possible) and/or discover (a) given sequence(s). These simple actions constitute a group of discrete movements that together form a more complex movement acquired through stimulus-response mapping (Krakauer et al., 2019). The main motor learning outcomes of SQTs are usually reaction time and accuracy, which are often combined to create a performance index. SQTs have been shown to engage brain regions and motor networks similar to those activated during SMTs but require a much higher involvement of cortical regions such as the left dorsal premotor cortex, supplementary motor cortex, and superior parietal cortex (Hardwick et al., 2013). These cortical regions are even more engaged when the SQTs have to be learned explicitly (Hardwick et al., 2013). Importantly, SQTs that implement higher-order sequence(s) also activate the medial temporal lobe, which effectively supports the higher temporal component of the task (Poldrack and Rodriguez, 2003; Robertson, 2007). Sequential movement using, for example, a response keypad, keyboard, or generic buttons, such as the serial reaction time task and its variations, as well as the $m \times n$ task are considered SQTs (Karni et al., 1995; Knowlton et al., 2017b; Krakauer et al., 2019). |
| **Visuomotor Adaptation Tasks (VATs)** require the learner to modify an already well-practiced motor behavior to respond to visual perturbations to maintain or regain the same levels of performance as before the introduction of the visual distortion. The motor system achieves this goal by updating existing motor programs rather than building entirely new motor routines. The main motor learning outcome of VATs is generally measured in form of directional error, defined as the angular difference between the target/goal movement and the actual movement. However, VATs can also involve changes in movement kinematics (e.g., reaction time and peak velocity). VATs have been shown to engage the cerebellum, basal ganglia (i.e., striatum), primary motor cortex, and fronto-parietal lobules as well as other sensorimotor-related cortical areas. Visuomotor tasks requiring to respond to a visual perturbation are categorized as VATs (Krakauer, 2009; Krakauer et al., 2005; Krakauer et al., 2019; Krakauer et al., 2000; Krakauer and Shadmehr, 2006). |
| **Gross Motor Tasks (GMTs)** involve the whole body and require the learner to maintain a stable position while performing different types of postural and balance tasks. Based on their characteristics and requirements (Dijkstra et al., 2020), these postural/balance tasks are categorized as static, dynamic and reactive. The main motor learning outcomes of GMTs vary considerably, but normally they measure time in balance, reaction time, variables associated with CoP/CoM displacement (e.g., CoP velocity), and the root mean square error (RMSE), i.e., in whole-body tracking tasks. GMTs have been shown to engage the brainstem, cerebellum, basal ganglia, thalamus, and several cortical areas within the temporal, parietal and frontal lobes (Dijkstra et al., 2020; Sargent et al., 2019) as well as different sensory systems (i.e., somatosensory, vestibular and visual systems) (Ivanenko and Gurfinkel, 2018). The frontal regions seem to play a critical role in GMTs, particularly in dynamic balance tasks, possibly supporting the development of task-specific strategies (Taubert et al., 2010). Gross postural/balance tasks involving the whole body were categorized as GMTs. |
| **Speech Motor Tasks (SPTs)** require the learner to build new motor commands to either produce novel nonsensical sequences of syllables or nonword phoneme sequences or to adapt already mastered speech motor schemes to efficiently respond to perturbations introduced in form of altered (auditory and/or somatosensory) feedback. SPTs can be classified as speech motor sequence learning or sensorimotor speech adaptation. Given that the (main) goal of speech is to produce acoustic or auditory signals that convey specific information (Guenther, 2016), the main motor learning outcomes investigated in SPT studies are generally utterance duration, error rate (i.e., accuracy), reaction time (Sternberg et al., 1978), sequencing errors (Masapollo et al., 2021), voice amplitude, fundamental frequency, and articulatory movements (e.g., lip speed) (Guenther, 2016). SPTs have been shown to engage several areas of the brain associated with working memory, speech motor planning, as well as the basal-ganglia-thalamocortical loop, and the cerebellum. SPTs seem to be associated with the integrity of structural connectivity between the motor and sensory brain regions (Alm, 2021; Frank et al., 2000; Maas et al., 2008; Segawa et al., 2015). |
| **Footnotes:** CoM=centre of mass; CoP=centre of pressure. |

When at least 10 effect sizes were available (Harrer et al., 2019), meta-regression was conducted to investigate the influence of moderators and their interactions. Moderators included different features of PD (Marinelli et al., 2017a) such as duration (i.e., years since diagnosis), severity of the disease (i.e., Hoehn and Yahr Score (Goetz et al., 2004) and the motor score of the Unified Parkinson’s Disease Rating Scale -UPDRS-Part III Motor Examination) (Goetz et al., 2008; Hentz et al., 2015; Hoehn and Yahr, 1967). The moderating effect of the methodological quality of the studies (**Suppl. 3)** and, when possible, the effect of anti-parkinsonian medication (“on” vs. “off”), were also investigated (**Suppl. 4**).

Considering that differences in motor skill acquisition between PD and NI groups (Clark et al., 2014; Felix et al., 2012; Marinelli et al., 2017a; Siegert et al., 2006) could influence retention, we also investigated its potential moderating effect using meta-regression and grouping studies that showed a significant improvement in skill acquisition during practice in favour of either NI individuals or PD patients, or that showed no difference between groups. We also explored deficits in skill transfer, which are reported separately in the supplementary files (**Suppl. 5**).

When a study used different variations of the same motor task that still required similar motor and cognitive demands and it was possible to calculate multiple effect sizes, we pooled them together by creating a composite Z-score (Higgins and Green, 2011). By contrast, when variations in the experimental conditions (e.g., blocked vs. random practice) were substantial, and thus potentially affecting the rate of acquisition and/or retention (Kantak et al., 2010; Schmidt et al., 2018), we did not create a composite Z-score and treated the different conditions separately (**Suppl. 6**). All analyses were conducted using the primary outcome of the motor tasks.

### 2.6. Data analysis

Analyses were performed with R (https://www.r-project.org; version 4.0.3) using the packages meta, metafor, ggplot2, and robvis (Harrer et al., 2019). Data entered for each group included mean differences and pooled SD (SDpooled) for different endpoints, as well as the number of participants in each group. Data were analyzed as continuous variables using a random-effects model, the restricted maximum likelihood (REML), and the “Hedges” procedure method, to calculate the standardized mean difference (SMD) with a 95% confidence interval (CI) (Harrer et al., 2019; Higgins et al., 2021).

We calculated the group mean difference in skill retention using end of practice and retention test scores (retention score – end of practice score) (Schmidt et al., 2018). If end of practice scores could not be obtained, we used the scores of the retention test performed immediately after the end of acquisition (Roig et al., 2013). Similarly, skill acquisition was calculated using the mean difference between scores obtained at baseline and the end of practice, either on a single session or multiple sessions (i.e., extended practice).

A p-value of less than 0.05 indicated statistical significance for an overall effect. Negative values represented worse skill retention scores for persons with PD in comparison with NI individuals. Heterogeneity was assessed with the I^2^ index, which was categorized as: low (0% to <40%), moderate (≥40% to <60%), substantial (≥60% to ≥75%) or large (≥75%) (Higgins and Green, 2011) and its statistical significance was assessed with the Cochran’s Q test (Harrer et al., 2019).

For meta-regression, we implemented the steps outlined by Harrer et al., (2019) (Harrer et al., 2019) and followed the guidelines provided by Veroniki and colleagues (Veroniki et al., 2016). Multicollinearity among predictors (*r* ≥ 0.8) (Harrer et al., 2019) was investigated using correlation matrices. To confirm the robustness of the meta-regression results and verify their true significance, we conducted permutation tests as described by Harrer et al., (2019) (Harrer et al., 2019). Permutation tests are a resampling method used to adjust the p-value of the meta-regression and thus control for type I error, which can be inflated when heterogeneity is present (Higgins and Thompson, 2004).

## 3. Results

### 3.1. Articles retrieved

The stages of the search and review processes with the main reasons for exclusion can be found in **Suppl. 7**. The electronic search yielded 9003 records but 59 additional studies from previous reviews were added. After removing duplicates, 3814 abstracts were screened with 185 studies reviewed at full-text level. Ninety-six studies were excluded because they did not have a retention test, or the latter was assessed less than one hour after the end of practice (see eligibility criteria). Nineteen studies were excluded because they did not use an appropriate motor task/method to assess motor learning and thirteen studies used a study design that did not meet the inclusion criteria. Two studies used deep brain stimulation, one study included a control group with neurological conditions, and another study had a control group with an age that differed significantly from the PD group. Four studies were excluded due to the lack of a control/PD group and four studies were excluded for other reasons (e.g., duplicated data). After identifying 45 studies, one additional study (Nutt et al., 2000) found in the reference list of studies reviewed was added. The review included a total of 46 studies but since it was not possible to obtain means and SDs from six studies (Doyon et al., 1998; Gawrys et al., 2008; Isaias et al., 2011; Nutt et al., 2000; ThomasAnterion et al., 1996; Werheid et al., 2003), whose results are reported qualitatively, the meta-analyses included 40 studies (Agostino et al., 2004; Behrman et al., 2000; Dan et al., 2015; Dantas et al., 2018; Foreman et al., 2013; Hadj-Bouziane et al., 2013; Harrington et al., 1990; Hayes and Hunsaker, 2015; Jessop et al., 2006; Kawashima et al., 2018; Lahlou et al., 2022; Lee et al., 2016; Lee and Fisher, 2018; Lee et al., 2019; Leow et al., 2012; Lin et al., 2007; Marinelli et al., 2009; Marinelli et al., 2017b; Mochizuki-Kawai et al., 2010; Nackaerts et al., 2020; Nelson et al., 2017; Nicastro et al., 2018; Onla-Or and Winstein, 2008; Pendt et al., 2012; Pendt et al., 2011; Peterson et al., 2016; Platz et al., 1998; Rostami and Ashayeri, 2009; Roy et al., 2015; Sato et al., 2014; Sehm et al., 2014; Sidaway et al., 2016; Simley-Oyen et al., 2002; Smiley-Oyen et al., 2012; Smiley-Oyen et al., 2006; Smiley-Oyen et al., 2003; Swinnen et al., 2000; Terpening et al., 2013; Van Ooteghem et al., 2017; Whitfield and Goberman, 2017).

### 3.2. Characteristics of the studies

A detailed summary of the 46 studies included in the review is reported in **Table 2**. Overall, data from 652 persons living with PD and 620 NI individuals acting as control were included. Of these participants, 550 were males and 423 were females, while the sex of 299 participants was not reported. The studies investigated mainly older adults with mean ages ranging from 52 to 74. Disease severity, which was not reported in eight studies (Agostino et al., 2004; Lee et al., 2019; Leow et al., 2012; Mochizuki-Kawai et al., 2010; Nutt et al., 2000; Roy et al., 2015; Simley-Oyen et al., 2002; Smiley-Oyen et al., 2003), ranged from I to IV on the Hoehn & Yahr scale. The mean overall scores of the motor evaluation conducted with the UPDRS part III, which was reported in 22 studies (Agostino et al., 2004; Dan et al., 2015; Hayes and Hunsaker, 2015; Isaias et al., 2011; Kawashima et al., 2018; Lahlou et al., 2022; Lee et al., 2019; Leow et al., 2012; Lin et al., 2007; Marinelli et al., 2017b; Nackaerts et al., 2020; Nelson et al., 2017; Nicastro et al., 2018; Pendt et al., 2012; Pendt et al., 2011; Peterson et al., 2016; Rostami and Ashayeri, 2009; Roy et al., 2015; Sehm et al., 2014; Sidaway et al., 2016; Van Ooteghem et al., 2017; Whitfield and Goberman, 2017), ranged from 8 to 33.4, indicating that the severity of motor symptoms of patients ranged from mild to moderate (Hentz et al., 2015; Martínez-Martín et al., 2015). Disease duration, which was reported in all but seven studies (Dantas et al., 2018; Marinelli et al., 2017b; Mochizuki-Kawai et al., 2010; Sato et al., 2014; Sidaway et al., 2016; Simley-Oyen et al., 2002; Smiley-Oyen et al., 2003), ranged from 1.3 to 11.1 years. Only three investigations (Hadj-Bouziane et al., 2013; Hayes and Hunsaker, 2015; Lahlou et al., 2022) manipulated (e.g., compared “on” vs. “off”) medication status, while two studies (Kawashima et al., 2018; Platz et al., 1998) tested patients “off” medication (**Table 2**). Six studies did not report and/or evaluate 10 cognitive functioning (Foreman et al., 2013; Hadj-Bouziane et al., 2013; Jessop et al., 2006; Nelson et al., 2017; Nutt et al., 2000; Van Ooteghem et al., 2017). In all, except five studies (Gawrys et al., 2008; Harrington et al., 1990; Hayes and Hunsaker, 2015; Simley-Oyen et al., 2002; Smiley-Oyen et al., 2003) that reported small differences in cognition between groups, PD and NI participants had similar cognitive status. None of the studies that explored associations between cognitive scores and skill acquisition or retention found significant correlations (Gawrys et al., 2008; Harrington et al., 1990; Nackaerts et al., 2020; Peterson et al., 2016; Sato et al., 2014).

**Table 2.**
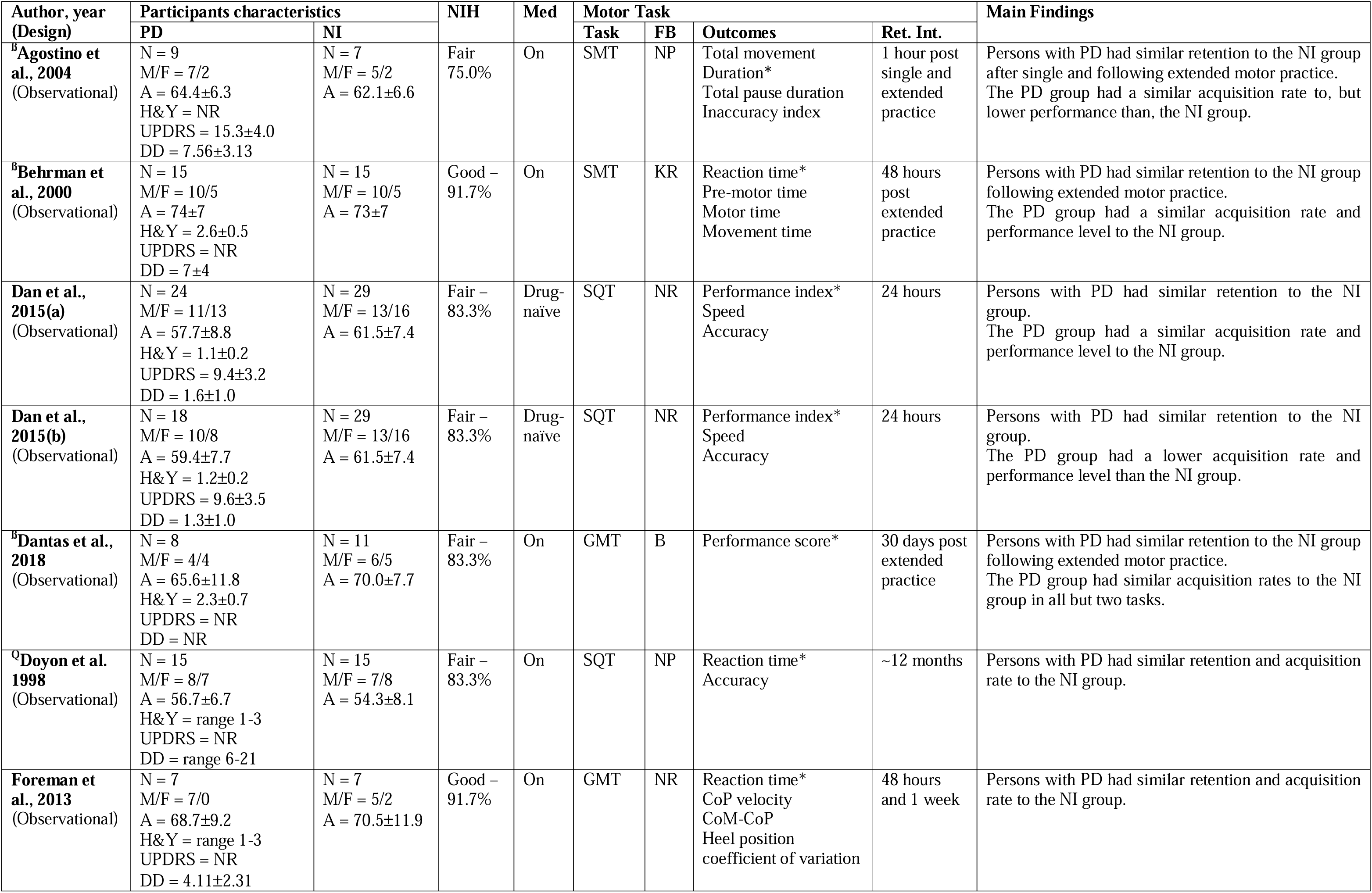
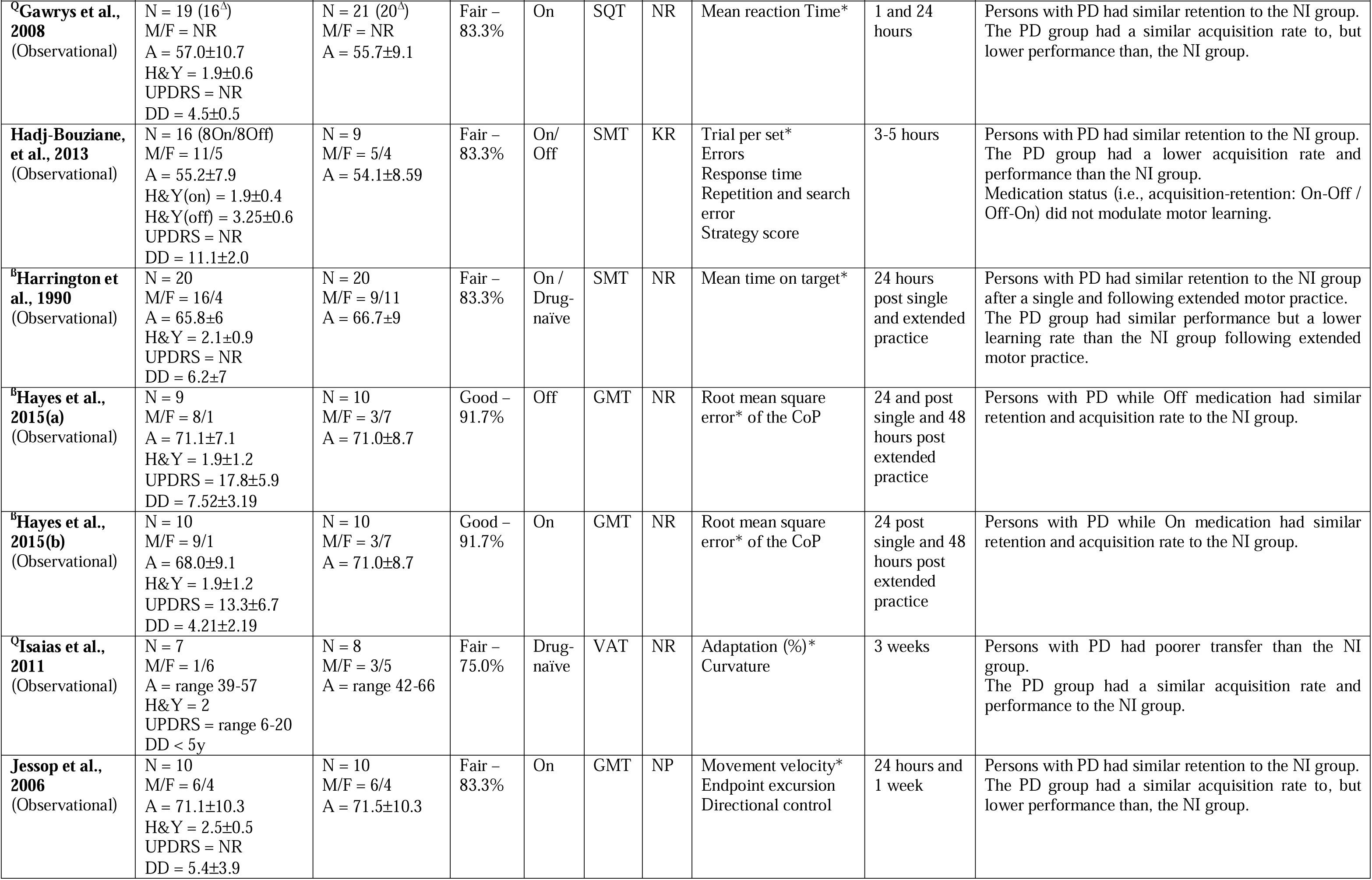
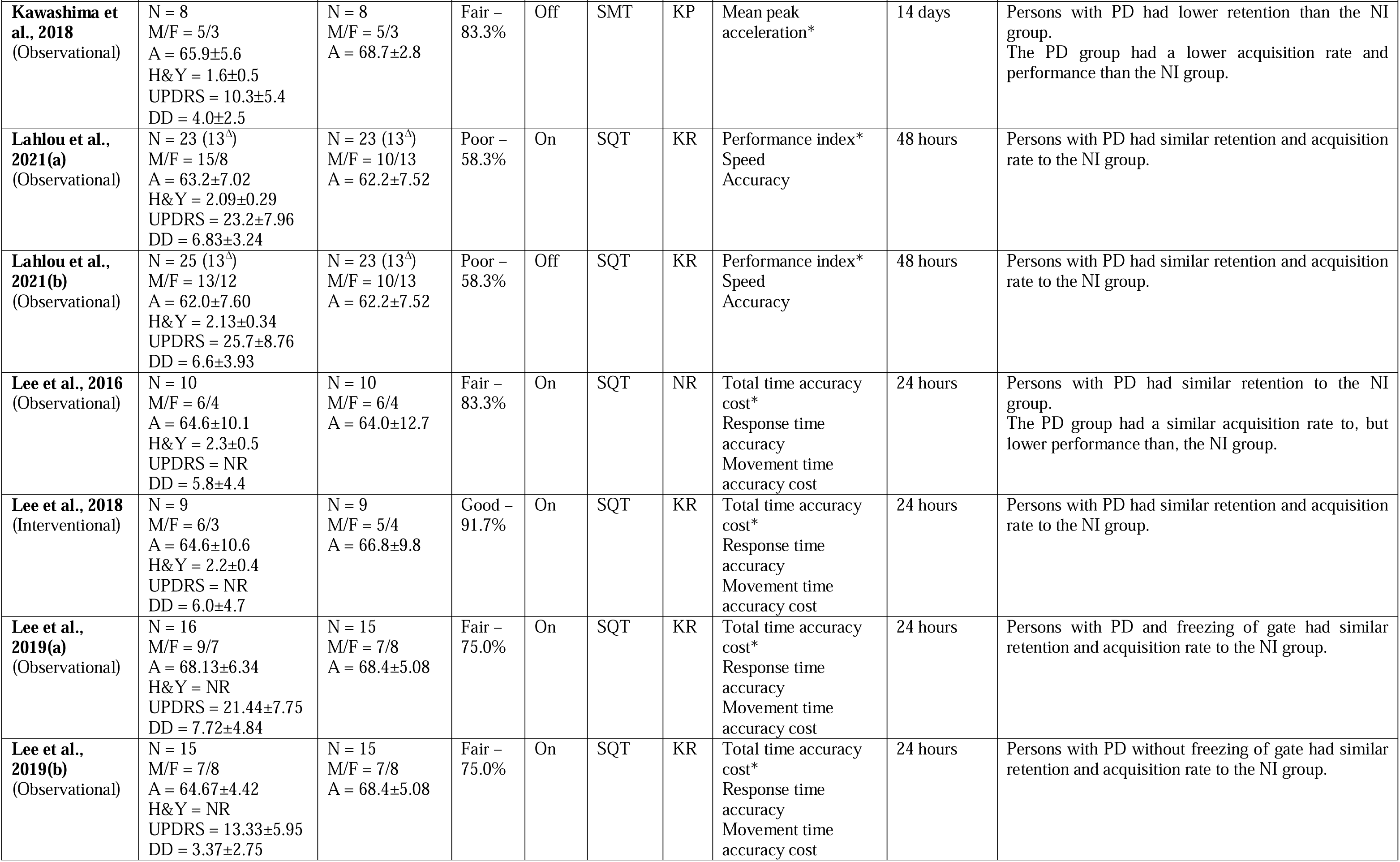
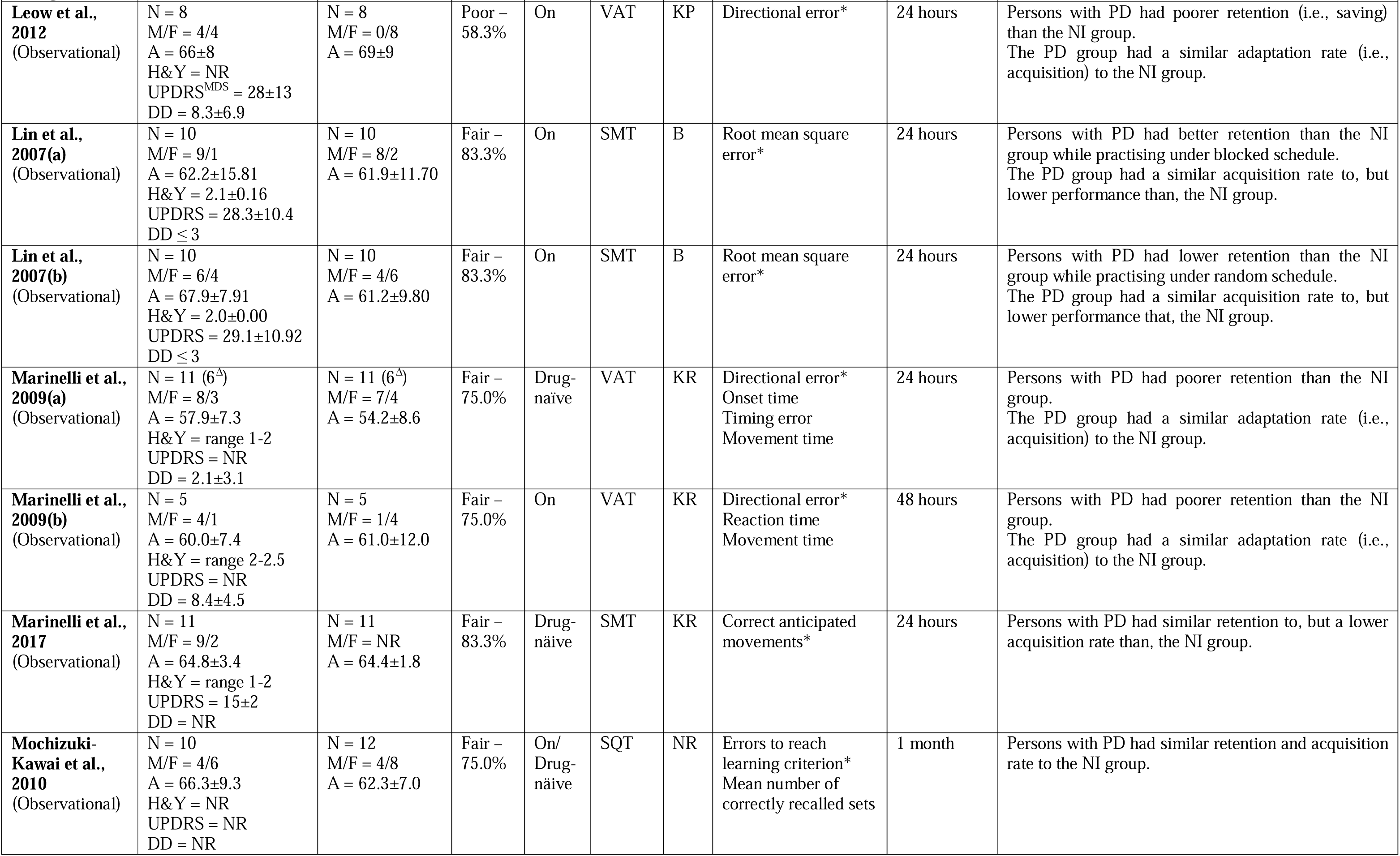
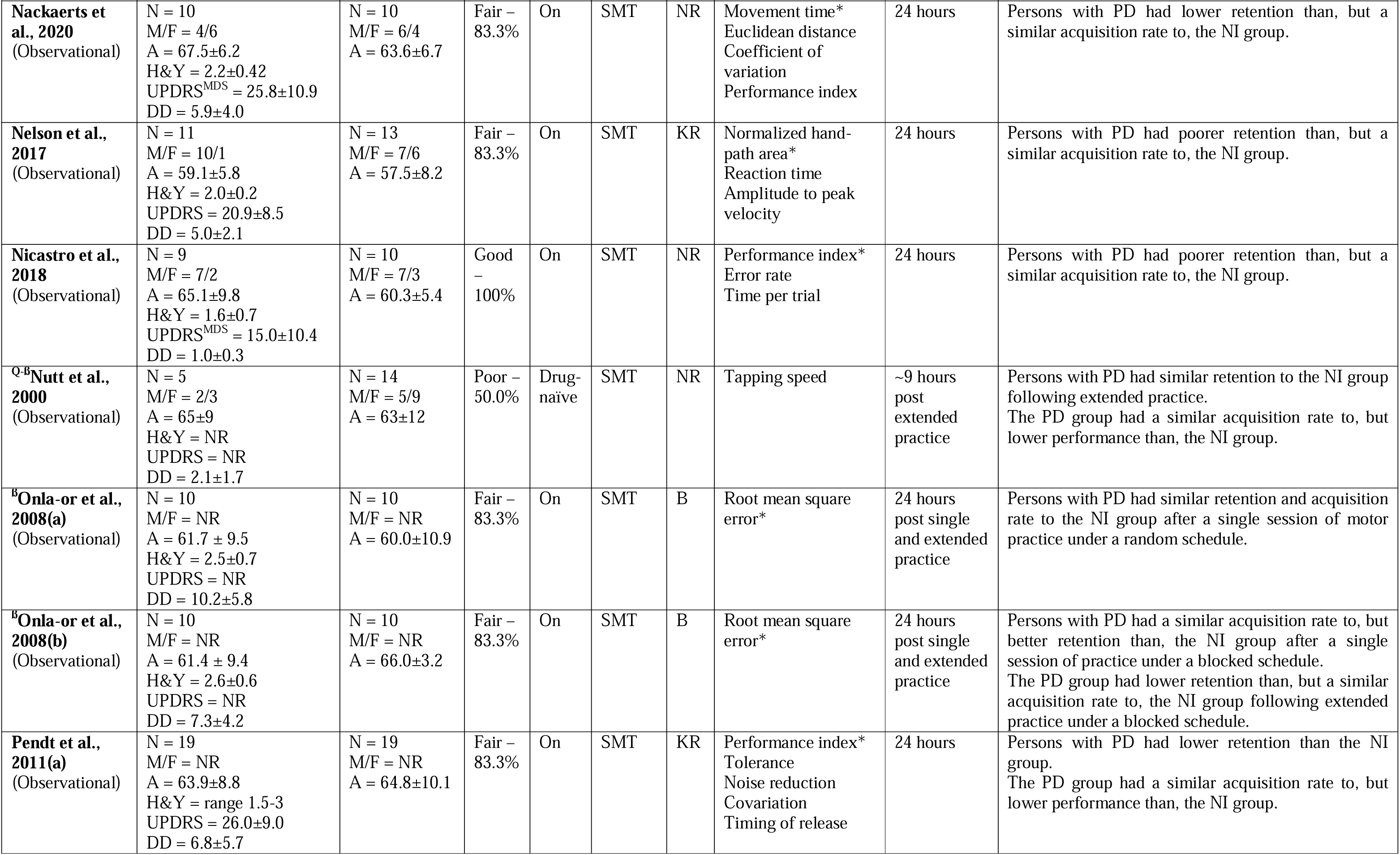
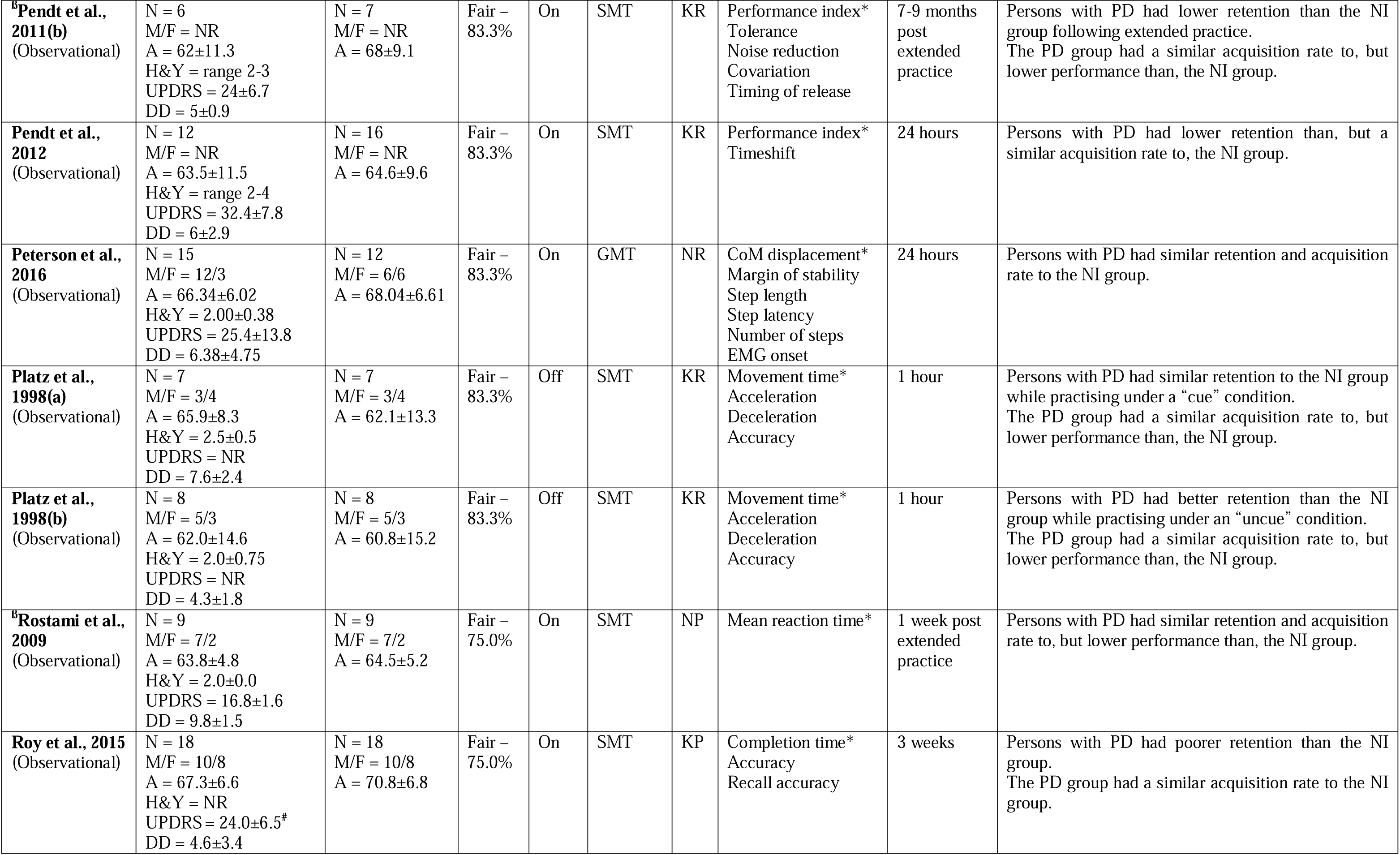
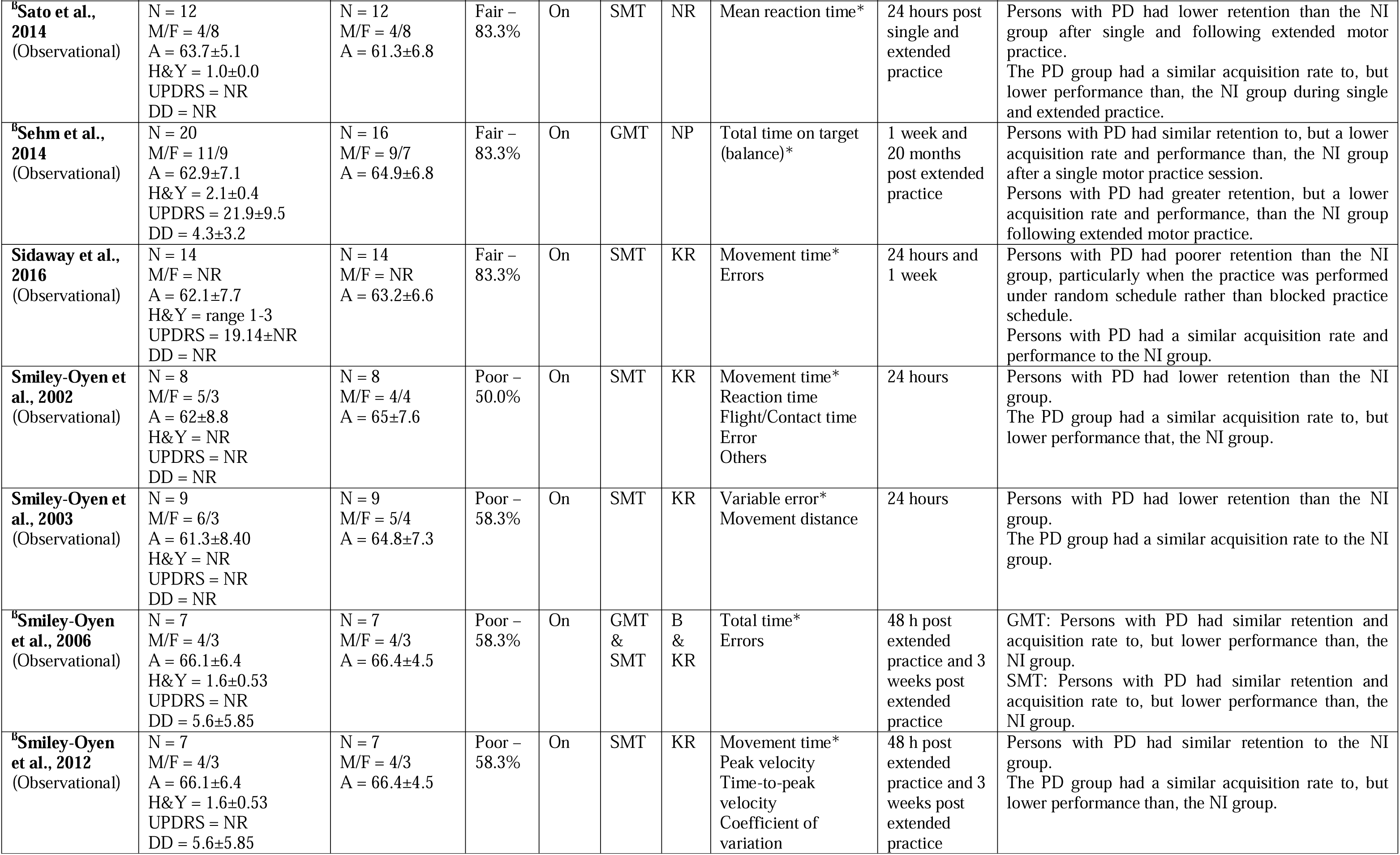
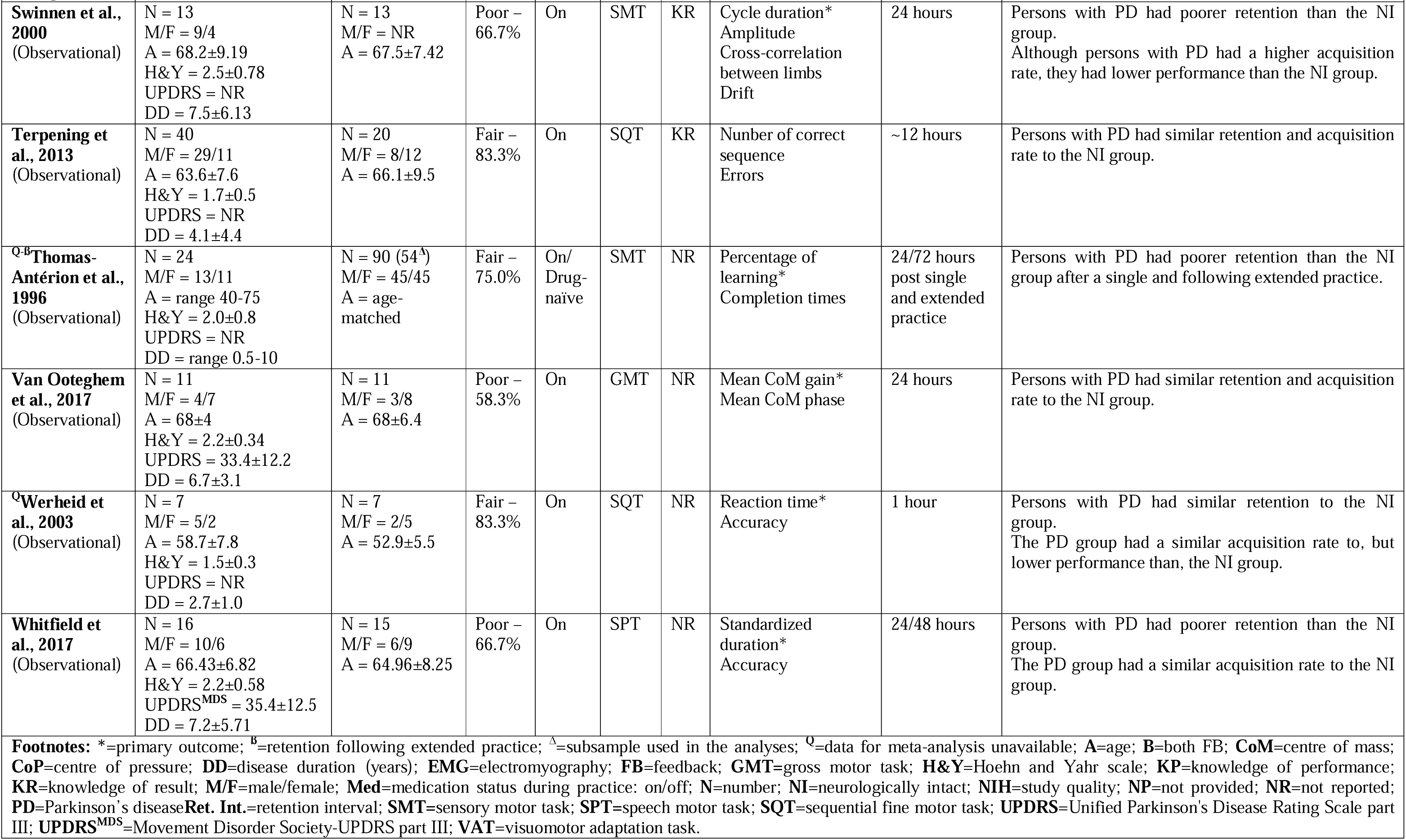
Study characteristics

Forty studies (Agostino et al., 2004; Dan et al., 2015; Doyon et al., 1998; Foreman et al., 2013; Gawrys et al., 2008; Hadj-Bouziane et al., 2013; Harrington et al., 1990; Hayes and Hunsaker, 2015; Isaias et al., 2011; Jessop et al., 2006; Kawashima et al., 2018; Lahlou et al., 2022; Lee et al., 2016; Lee and Fisher, 2018; Lee et al., 2019; Leow et al., 2012; Lin et al., 2007; Marinelli et al., 2009; Marinelli et al., 2017b; Mochizuki-Kawai et al., 2010; Nackaerts et al., 2020; Nelson et al., 2017; Nicastro et al., 2018; Onla-Or and Winstein, 2008; Pendt et al., 2012; Pendt et al., 2011; Peterson et al., 2016; Platz et al., 1998; Roy et al., 2015; Sato et al., 2014; Sehm et al., 2014; Sidaway et al., 2016; Simley-Oyen et al., 2002; Smiley-Oyen et al., 2003; Swinnen et al., 2000; Terpening et al., 2013; ThomasAnterion et al., 1996; Van Ooteghem et al., 2017; Werheid et al., 2003; Whitfield and Goberman, 2017) assessed retention after a single session of practice, while 14 studies (Agostino et al., 2004; Behrman et al., 2000; Dantas et al., 2018; Harrington et al., 1990; Hayes and Hunsaker, 2015; Nutt et al., 2000; Onla-Or and Winstein, 2008; Pendt et al., 2011; Rostami and Ashayeri, 2009; Sato et al., 2014; Sehm et al., 2014; Smiley-Oyen et al., 2012; Smiley-Oyen et al., 2006; ThomasAnterion et al., 1996) investigated retention after extended practice (≥2 sessions). Regarding the nature of the task, 25 studies used SMTs (Agostino et al., 2004; Behrman et al., 2000; Hadj-Bouziane et al., 2013; Harrington et al., 1990; Kawashima et al., 2018; Lin et al., 2007; Marinelli et al., 2017b; Nackaerts et al., 2020; Nelson et al., 2017; Nicastro et al., 11 2018; Nutt et al., 2000; Onla-Or and Winstein, 2008; Pendt et al., 2012; Pendt et al., 2011; Platz et al., 1998; Rostami and Ashayeri, 2009; Roy et al., 2015; Sato et al., 2014; Sidaway et al., 2016; Simley-Oyen et al., 2002; Smiley-Oyen et al., 2012; Smiley-Oyen et al., 2006; Smiley-Oyen et al., 2003; Swinnen et al., 2000; ThomasAnterion et al., 1996); 10 studies SQTs (Dan et al., 2015; Doyon et al., 1998; Gawrys et al., 2008; Lahlou et al., 2022; Lee et al., 2016; Lee and Fisher, 2018; Lee et al., 2019; Mochizuki-Kawai et al., 2010; Terpening et al., 2013; Werheid et al., 2003); three studies VATs (Isaias et al., 2011; Leow et al., 2012; Marinelli et al., 2009); eight studies GMTs (Dantas et al., 2018; Foreman et al., 2013; Hayes and Hunsaker, 2015; Jessop et al., 2006; Peterson et al., 2016; Sehm et al., 2014; Smiley-Oyen et al., 2006; Van Ooteghem et al., 2017), and one study SPTs (Whitfield and Goberman, 2017). Nineteen studies (Behrman et al., 2000; Hadj-Bouziane et al., 2013; Isaias et al., 2011; Lahlou et al., 2022; Lee and Fisher, 2018; Lee et al., 2019; Marinelli et al., 2009; Marinelli et al., 2017b; Nelson et al., 2017; Pendt et al., 2012; Pendt et al., 2011; Platz et al., 1998; Sidaway et al., 2016; Simley-Oyen et al., 2002; Smiley-Oyen et al., 2012; Smiley-Oyen et al., 2006; Smiley-Oyen et al., 2003; Swinnen et al., 2000; Terpening et al., 2013) provided feedback in the form of knowledge of results (e.g., numeric score), three (Kawashima et al., 2018; Leow et al., 2012; Roy et al., 2015) as knowledge of performance (e.g., movement trajectory), and four (Dantas et al., 2018; Lin et al., 2007; Onla-Or and Winstein, 2008; Smiley-Oyen et al., 2006) combined these two types of feedback. Of the remaining studies, five studies (Agostino et al., 2004; Doyon et al., 1998; Jessop et al., 2006; Rostami and Ashayeri, 2009; Sehm et al., 2014) did not provide feedback and 16 (Dan et al., 2015; Foreman et al., 2013; Gawrys et al., 2008; Harrington et al., 1990; Hayes and Hunsaker, 2015; Lee et al., 2016; Mochizuki-Kawai et al., 2010; Nackaerts et al., 2020; Nicastro et al., 2018; Nutt et al., 2000; Peterson et al., 2016; Sato et al., 2014; ThomasAnterion et al., 1996; Van Ooteghem et al., 2017; Werheid et al., 2003; Whitfield and Goberman, 2017) did not explicitly state if feedback was provided or not.

### 3.3. Methodological quality

Details of the methodological quality assessment can be found in **Suppl. 8**. The percentage score and quality rating of each study are reported in **Table 2**. The mean±SD percentage score was 77.5±11.6% with 31 studies rated as “fair”, five as “good” and 10 as “poor”. The most common methodological flaws were the lack of both a sample size justification and information regarding the number of participants who were excluded from participation because they did not meet the inclusion criteria. The visual inspection of the funnel plots (**Suppl. 9**) and the results from Egger’s regressions (**Suppl. 10)** including studies of the meta-analyses, suggested no, or minimal, presence of heterogeneity and risk of biases at the study level. Finally, meta-regressions using the methodological quality as a covariate showed non-significant results, suggesting that low quality studies did not inflate between-group differences in skill retention after single (*p* = 0.1750) (**Suppl. 3**) or extended practice (*p* = 0.6850).

### 3.4. Retention after a single practice session

Overall, 17 (Isaias et al., 2011; Kawashima et al., 2018; Leow et al., 2012; Marinelli et al., 2009; Nackaerts et al., 2020; Nelson et al., 2017; Nicastro et al., 2018; Pendt et al., 2012; Pendt et al., 2011; Roy et al., 2015; Sato et al., 2014; Sidaway et al., 2016; Simley-Oyen et al., 2002; Smiley-Oyen et al., 2003; Swinnen et al., 2000; ThomasAnterion et al., 1996; Whitfield and Goberman, 2017) of the 40 studies that investigated skill retention after a single practice session (42.5%), reported that persons with PD had poorer skill retention than the control group. These 17 studies employed SMTs, VATs, and SPTs (**Table 2**). The remaining 23 studies, most of which implemented SQTs and GMTs, revealed no significant differences in skill retention between groups (Agostino et al., 2004; Dan et al., 2015; Doyon et al., 1998; Foreman et al., 2013; Gawrys et al., 2008; Hadj-Bouziane et al., 2013; Harrington et al., 1990; Hayes and Hunsaker, 2015; Jessop et al., 2006; Lahlou et al., 2022; Lee et al., 2016; Lee and Fisher, 2018; Lee et al., 2019; Lin et al., 2007; Marinelli et al., 2017b; Mochizuki-Kawai et al., 2010; Onla-Or and Winstein, 2008; Peterson et al., 2016; Platz et al., 1998; Sehm et al., 2014; Terpening et al., 2013; Van Ooteghem et al., 2017; Werheid et al., 2003).

When pooled together in the meta-analysis, data from the 35 studies (Agostino et al., 2004; Dan et al., 2015; Foreman et al., 2013; Hadj-Bouziane et al., 2013; Harrington et al., 1990; Hayes and Hunsaker, 2015; Jessop et al., 2006; Kawashima et al., 2018; Lahlou et al., 2022; Lee et al., 2016; Lee and Fisher, 2018; Lee et al., 2019; Leow et al., 2012; Lin et al., 2007; Marinelli et al., 2009; Marinelli et al., 2017b; Mochizuki-Kawai et al., 2010; Nackaerts et al., 2020; Nelson et al., 2017; Nicastro et al., 2018; Onla-Or and Winstein, 2008; Pendt et al., 2012; Pendt et al., 2011; Peterson et al., 2016; Platz et al., 1998; Roy et al., 2015; Sato et al., 2014; Sehm et al., 2014; Sidaway et al., 2016; Simley-Oyen et al., 2002; Smiley-Oyen et al., 2003; Swinnen et al., 2000; Terpening et al., 2013; Van Ooteghem et al., 2017; Whitfield and Goberman, 2017) (47 effect sizes and 1187 participants) investigating skill retention after a single practice session showed a small significant effect in favor of NI individuals (SMD = −0.17; 95% CI = −0.32, −0.02; *p* = 0.0225; N = 47; I^2^ = 39.6%). Heterogeneity was low but statistically significant (Q-test: *p* = 0.0034). Sub-group analyses revealed a significant moderating effect of the task nature (*p* < 0.0001) (**Figure 1**) and type of feedback (*p* = 0.0238) (**Table 3**). Specifically, people with PD showed worse skill retention in both SMTs (SMD = −0.31; 95% CI −0.47, −0.15; N = 25; I^2^ = 27.4%; *p* = 0.0002) and VATs (SMD = − 1.55; 95% CI = −2.32, −0.79; N = 3; I^2^ = 0%; *p* = 0.0001) but not in SQTs (SMD = 0.17; 95% CI = −0.05, 0.39; N = 10; I^2^ = 0%; *p* = 0.1292) or GMTs (SMD = 0.04; 95% CI = −0.25, 0.33; N = 8; I^2^ = 0%; *p* = 0.7771). The only study that investigated SPTs showed a large effect in favour of the NI group (SMD = −1.28; 95% CI = −2.07, −0.50; *p* = 0.0013). Sub-group analyses pertaining to feedback are reported in **Table 3**.

**Figure 1.**
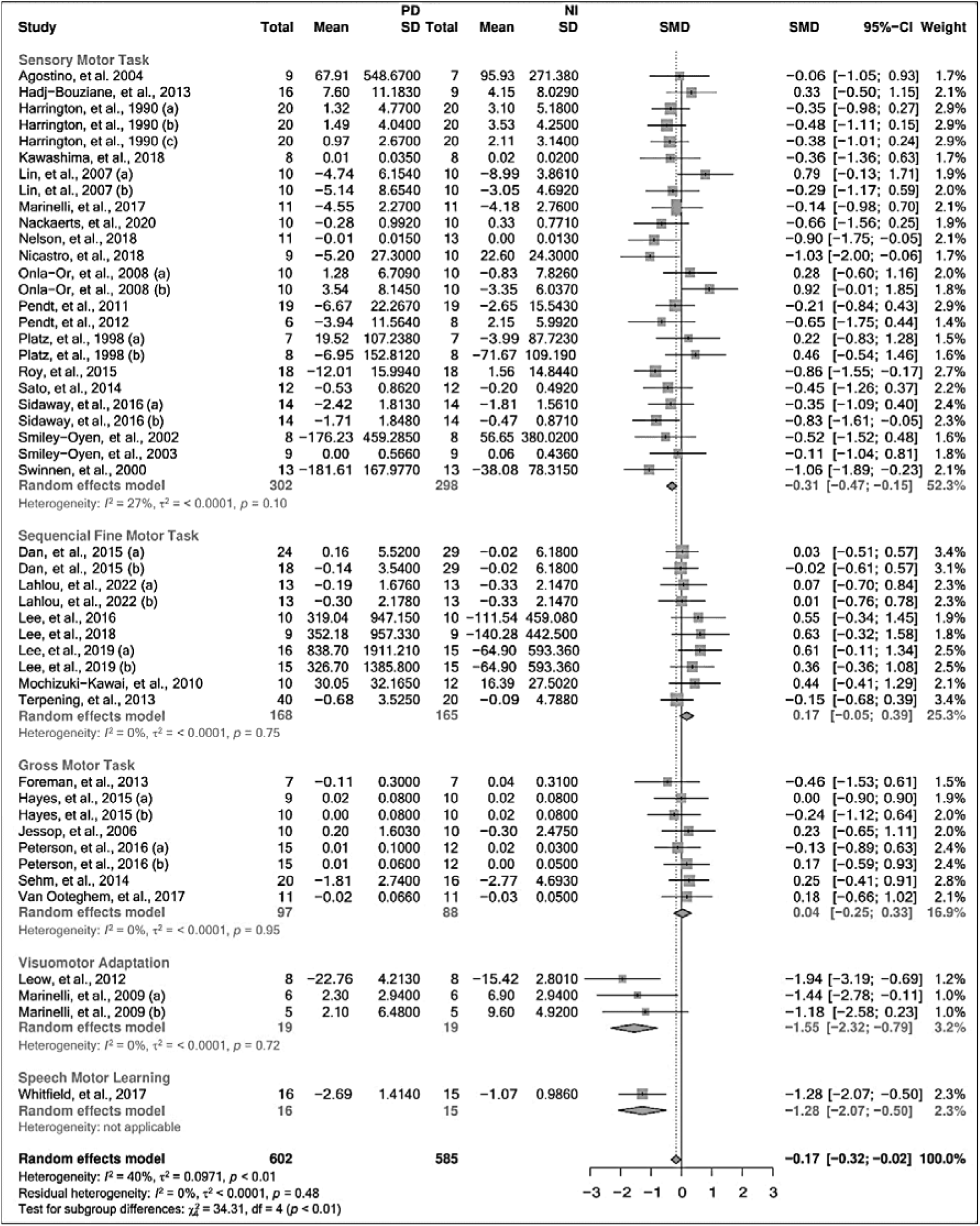
Motor skill retention after a single session of practice grouped by the task nature; CI = confidence interval; NI = neurologically intact individuals; PD = people with Parkinson’s Disease; SD = standard deviation; SMD = standard mean difference.

**Table 3.**
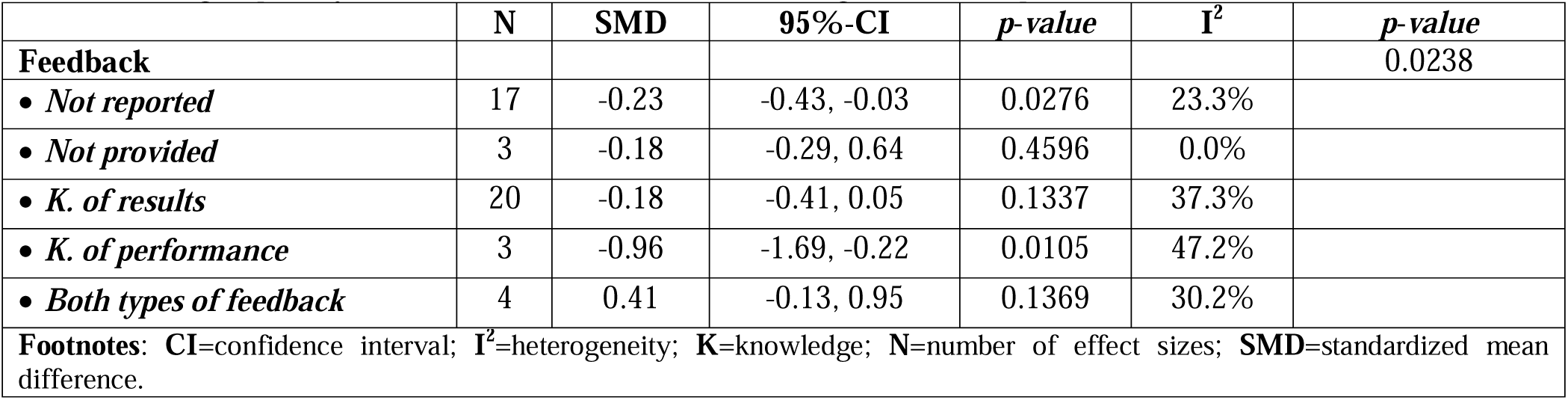
Sub-group analysis for motor skill retention after a single session of practice.

|  | **N** | **SMD** | **95%-CI** | ***p-value*** | **I<sup>2</sup>** | ***p-value*** |
| --- | --- | --- | --- | --- | --- | --- |
| **Feedback** |  |  |  |  |  | 0.0238 |
| • *Not reported* | 17 | -0.23 | -0.43, -0.03 | 0.0276 | 23.3% |  |
| • *Not provided* | 3 | -0.18 | -0.29, 0.64 | 0.4596 | 0.0% |  |
| • *K. of results* | 20 | -0.18 | -0.41, 0.05 | 0.1337 | 37.3% |  |
| • *K. of performance* | 3 | -0.96 | -1.69, -0.22 | 0.0105 | 47.2% |  |
| • *Both types of feedback* | 4 | 0.41 | -0.13, 0.95 | 0.1369 | 30.2% |  |
| **Footnotes:** CI=confidence interval; I<sup>2</sup>=heterogeneity; K=knowledge; N=number of effect sizes; SMD=standardized mean difference. |  |  |  |  |  |  |

The results of the meta-regression exploring the influence of disease duration and severity, as well as their interactions are reported in **Suppl. 3**. None of these analyses yielded significant results, suggesting that disease duration and severity did not affect motor skill retention following a single session of practice. Similarly, skill retention did not seem to be moderated by improvements in skill performance during acquisition (**Suppl. 3**). Due to the small number of studies, we could not establish direct comparisons to investigate potential mediating effects of medication status (“on” vs. “off”). However, sensitivity analyses revealed that studies that manipulated medication status did not influence the results of the meta-analyses (**Suppl. 4)**.

### 3.5. Retention after extended practice

Only studies employing SMTs and GMTs investigated the effect of extensive practice on skill retention in PD. Therefore, the results pertaining to extended practice cannot be generalized to other types of motor tasks. Only two (Sato et al., 2014; ThomasAnterion et al., 1996) of the 14 studies that investigated skill retention following extended practice (14.3%) found that persons with PD had poorer skill retention than the control group. Except for one study (Sehm et al., 2014) that observed better retention in persons with PD using a GMT, the remaining studies reported no significant differences in skill retention between groups, regardless of whether SMTs (Agostino et al., 2004; Behrman et al., 2000; Harrington et al., 1990; Nutt et al., 2000; Onla-Or and Winstein, 2008; Pendt et al., 2011; Rostami and Ashayeri, 2009; Smiley-Oyen et al., 2012; Smiley-Oyen et al., 2006) or GMTs (Dantas et al., 2018; Hayes and Hunsaker, 2015; Smiley-Oyen et al., 2006) were implemented.

When pooled together in the meta-analysis, the data of 12 studies (Agostino et al., 2004; Behrman et al., 2000; Dantas et al., 2018; Harrington et al., 1990; Hayes and Hunsaker, 2015; Onla-Or and Winstein, 2008; Pendt et al., 2011; Rostami and Ashayeri, 2009; Sato et al., 2014; Sehm et al., 2014; Smiley-Oyen et al., 2012; Smiley-Oyen et al., 2006) studies (17 effect sizes and 403 participants) that investigated skill retention following extended practice using SMTs and GMTs revealed no significant differences between groups (SMD = −0.04; 95% CI = −0.24, 0.15; *p* = 0.6565; N = 17; I^2^ = 0%). Similarly, subgroup analyses did not reveal any significant moderating effect regarding the task nature (*p* = 0.0745) (**Figure 2**), or feedback provided (*p* = 0.9511).

**Figure 2.**
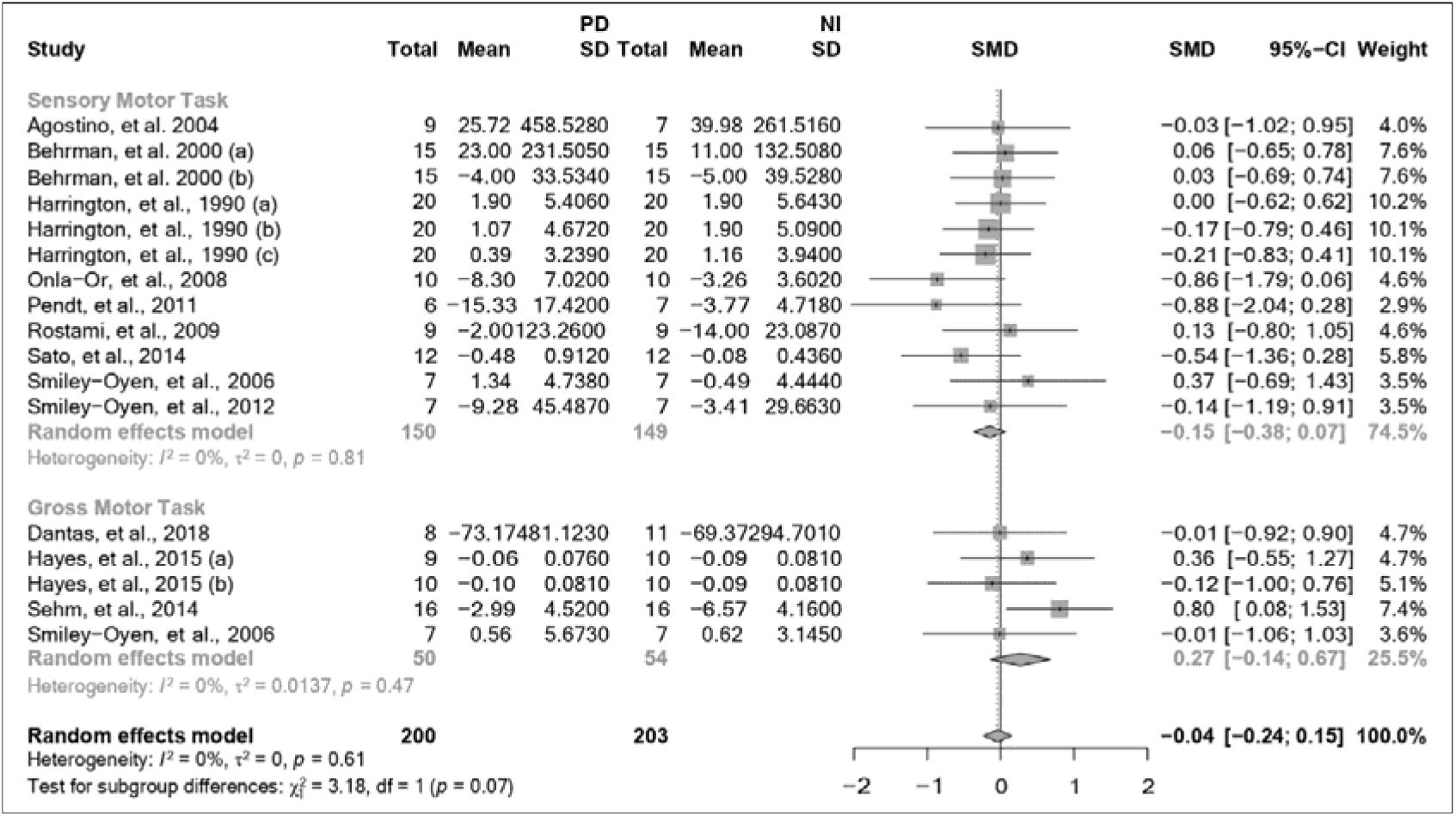
Motor skill retention after extended practice grouped by the task nature; CI = confidence interval; NI = neurologically intact individuals; PD = people with Parkinson’s Disease; SD = standard deviation; SMD = standard mean difference.

The results of meta-regression analyses exploring the influence of disease duration (*p* = 0.1980) and severity (*p* = 0.7730), as well as their interaction (*p* = 0.2600), were non-significant, indicating that these factors did not moderate skill retention after extended practice. Importantly, performance during skill acquisition did not appear to moderate retention either (*p* = 0.7162). Consistent with the results of single practice studies, sensitivity analyses showed that medication status did not influence the results of the meta-analyses including extended practice studies (**Suppl. 4).**

## 4. Discussion

The results of this review confirm that, in comparison with NI individuals of similar age, persons with mild to moderate PD have deficits in the capacity to retain motor skills after a single practice session. These findings, which have important clinical implications, suggest that to maximize the long-term retention of motor skills, motor rehabilitation should not only focus on optimizing skill acquisition and motor memory encoding but also on ensuring an effective motor memory consolidation process (Doyon, 2008; Marinelli et al., 2017a). Deficits in skill retention, however, do not affect all types of motor tasks to the same extent. Differences between PD and NI individuals were found to be statistically significant only in the retention of motor skills acquired during STMs, VATs and SPTs, although only one study investigated SPTs and thus these results should be interpreted cautiously. Patients with PD, in contrast, appear to have a more preserved capacity for retaining skills acquired during SQTs and GMTs. Motor rehabilitation programs aimed at maintaining motor function in PD should target a broad range of tasks to ensure functional mobility in multiple activities of daily living (Ferrazzoli et al., 2018; Nieuwboer et al., 2009). However, identifying task-specific deficits could inform clinicians to design more targeted interventions to optimize the acquisition and long-term retention of motor skills (Abbruzzese et al., 2016).

During the first symptomatic stages of PD (Blesa et al., 2022; Hawkes et al., 2010), neurodegenerative changes are localized within the basal ganglia and cortico-striatal motor networks while neocortical areas are less affected (Kordower et al., 2013; Obeso et al., 2008; Rodriguez-Oroz et al., 2009). Subsequently, structural and functional signs of deterioration appear in the cerebellum (Schindlbeck and Eidelberg, 2018; Wu and Hallett, 2013) and, to a lesser extent, cortical areas such as parietal (Tahmasian et al., 2017) and primary motor (M1) cortices (Ammann et al., 2020; Lindenbach and Bishop, 2013). As the disease progresses, neurodegeneration changes within the basal ganglia spread across the striatum (Kish et al., 1988; Kordower et al., 2013), affecting non-motor cortico-striatal circuits (Kehagia et al., 2013; Obeso et al., 2008; Rodriguez-Oroz et al., 2009) and the less affected contralateral side (Blesa et al., 2022; Pineda-Pardo et al., 2022). In the late stages of PD, neurodegeneration will eventually reach neocortical areas (Agosta et al., 2013; Hawkes et al., 2010). Whereas the acquisition and retention of SMTs involve mostly basal ganglia, cerebellum and cortical areas such as M1 (Doyon et al., 2009b; Hardwick et al., 2013; Hikosaka et al., 2002), VATs engage primarily the striatum (Huang et al., 2011; Krakauer et al., 2019), cerebellum, fronto-parietal lobules and M1 (Doyon et al., 2009b; Galea et al., 2011; Hardwick et al., 2013; Huber et al., 2004; Krakauer et al., 2019; Moisello et al., 2015) (**Table 1**). Neuroimaging studies have demonstrated that deficits in the retention of these types of motor tasks in PD are associated with alterations in parietal and cortico-striatal connectivity and dopamine uptake (Isaias et al., 2011; Kawashima et al., 2018; Manuel et al., 2018; Nelson et al., 2017; Nicastro et al., 2018). Deficits in the retention of motor skills acquired during the practice of SMTs and VATs in persons with mild to moderate PD could therefore be explained by the deterioration that these specific areas of the brain experience during the early symptomatic stages of the disease.

Given the broad implication of cortico-striatal networks in the acquisition of SQTs (Doyon et al., 2009b; Hikosaka et al., 2002; Lehéricy et al., 2005), the preserved capacity that patients with PD showed for retaining motor skills acquired during the practice of these motor tasks was unexpected. The reason for this preservation is unknown but it could be related to the capacity to activate neocortical areas that are consistently engaged during the practice of SQTs (Hardwick et al., 2013) and tend to be less affected by neurodegeneration in the early symptomatic stages of the disease (Hawkes et al., 2010). Indeed, compared to SMTs and VATs, the performance of SQTs is characterized by greater activation of cortical areas such as the left dorsal premotor cortex, the supplementary motor cortex as well as the superior parietal cortex (Hardwick et al., 2013). Increased participation of the cerebellum during motor practice could also contribute to the preservation of SQTs in people with PD (Appel-Cresswell et al., 2010; Mentis et al., 2003a; Mentis et al., 2003b; Simioni et al., 2016), although this brain structure displays signs of structural and functional alterations already during the Hoehn & Yahr stage II-III of the disease (Agosta et al., 2013; O’Callaghan et al., 2016). It is noteworthy that the activation of cortical and cerebellar regions is even more pronounced during the acquisition of explicit variants of the SQTs like the ones used in all the studies of the meta-analysis, in which participants are consciously aware of the repeating numerical sequence embedded in the motor task (Hardwick et al., 2013). Importantly, these patterns of brain activation are not only present during acquisition but also during motor memory consolidation (Sami et al., 2014). Clearly, more studies are needed to identify which mechanisms underlie the preserved capacity to retain SQTs shown by people with PD.

Deficits in the retention of SQTs could nevertheless become more pronounced when neurodegeneration progresses (Carbon et al., 2007; Carbon et al., 2010) and alterations in motor automaticity mechanisms, which are more relevant in the late phases of sequential motor learning, start to emerge (Redgrave et al., 2010; Wu et al., 2015). In one study included in the review, participants performed a retention test of a SQT 10–18 months after initial practice (Doyon et al., 1998) and only those patients whose disease severity worsened, transitioning from Hoehn and Yahr stage I to II (Hoehn and Yahr, 1967) during the retention interval, showed significant deficits in skill retention (Doyon et al., 1998). In addition, two studies of the review reported skill retention deficits only at the end of the second day of practice (Dan et al., 2015; Terpening et al., 2013), reinforcing the idea that deficits in motor skills practiced during SQTs become more pronounced in late phases of motor learning (Karni et al., 1998b), when sequential finger movements should be performed automatically and with less attentional demand. These results suggest that while patients in the initial symptomatic stages of the disease can acquire and retain sequential fine motor skills similarly to NI individuals, they can show deficits in the ability to automatize these motor skills (Redgrave et al., 2010; Wu et al., 2015). Cognitive strategies such as verbal instructions, cueing, and segmentation, could help those individuals to shift learning toward a more volitional mode of action to compensate for deficits in automaticity (Ferrazzoli et al., 2018).

The performance of GMTs requires complex postural control strategies involving multiple brain structures such as the brainstem, cerebellum, basal ganglia, thalamus, and neocortical areas (e.g., sensory-motor cortex), as well as the integration of information from proprioceptive, vestibular, and visual systems (Dijkstra et al., 2020; Ivanenko and Gurfinkel, 2018; Schoneburg et al., 2013). We found that people with mild to moderate PD can improve the performance of GMTs and retain these gains similarly to NI individuals. This finding is encouraging because patients with PD suffer from gait disorders and postural stability problems (Kim et al., 2013) that aggravate despite pharmacological treatment (Klawans, 1986; Mak et al., 2017) and they also fall significantly more so than NI individuals (Allen et al., 2013; Bloem et al., 2001; Boonstra et al., 2008; Canning et al., 2014; Stolze et al., 2005). GMT-based training could be a good strategy to slow down the deterioration of these complex motor skills (Abbruzzese et al., 2016) as this type of training can potentially mitigate alterations in neuroplasticity commonly observed in patients with PD (Zhuang et al., 2013). For example, step training increases intra-cortical inhibition measured with transcranial magnetic stimulation (Liu et al., 2022), an indirect marker of γ-aminobutyric acid (GABA) activity, which tends to be suppressed in people with PD (Ammann et al., 2020; Blesa et al., 2017; Rothwell and Edwards, 2013). Additionally, dynamic balance training has been shown to increase gray matter in the cerebellum, parietal and temporal lobes, as well as in the pre-motor cortex in patients with PD (Sehm et al., 2014). Taken together, these studies reinforce the importance of gait and postural control training in PD. Initiating these interventions as early as possible could potentially preserve neuroplasticity and slow down the progressive motor deterioration of the disease (Abbruzzese et al., 2016).

We used subgroup analyses and meta-regression techniques to explore factors moderating the capacity to retain motor skills in PD such as performance during acquisition, disease severity and duration, amount of practice and feedback provided. Although we cannot rule out the possibility that a ceiling effect in motor skill acquisition in NI individuals could have masked differences (Schmidt et al., 2018), the rate of skill improvement during acquisition was similar between groups. Regardless, acquisition performance did not appear to modulate the capacity to improve motor skill retention in patients with PD (**Suppl. 3**). Similarly, neither disease severity, disease duration, nor their interaction, had any significant influence on the observed deficits in motor skill retention, challenging previous studies indicating that these factors had detrimental effects on skill retention in people with PD (Carbon et al., 2007; Carbon et al., 2010; Clark et al., 2014; Doyon et al., 1998; Gawrys et al., 2008; Hadj-Bouziane et al., 2013; Harrington et al., 1990; Mochizuki-Kawai et al., 2010; Muslimovic et al., 2007; Nackaerts et al., 2020; Smiley-Oyen et al., 2006; Stephan et al., 2011; ThomasAnterion et al., 1996; Van Ooteghem et al., 2017). Discrepancies with previous studies could be explained by the fact that most studies included in our review recruited patients with mild-to-moderate PD. This possibly resulted in a very homogenous sample that limited the capacity of the meta-regression to capture moderating effects of disease severity or, to a lesser extent, disease duration on skill retention. Further research is needed to determine whether the capacity to retain some specific motor skills is affected or worsens more rapidly as the disease progresses.

Importantly, subgroup analyses revealed that deficits in the retention of motor skills were reduced when more practice was afforded (Marinelli et al., 2017b; Nieuwboer et al., 2009) and/or when augmented feedback was provided (Abbruzzese et al., 2016; Chiviacowsky et al., 2010; Marinelli et al., 2017a; Nieuwboer et al., 2009). With additional practice, patients with PD appear to be able to retain motor skills acquired during SMTs similarly to NI individuals (Abbruzzese et al., 2016; Ghilardi et al., 2003; Nieuwboer et al., 2009; Olson et al., 2019). Extended motor practice possibly allows patients the time to encode sensory and motor information more effectively, compensating for some of the neural deficits that they display during motor learning (Aslan et al., 2021; Nackaerts et al., 2019; Nelson et al., 2017). This is consistent with studies showing that, compared to NI individuals, persons in the early stages of PD require greater neural activity during practice to achieve similar skill performance (Aslan et al., 2021; Carbon and Eidelberg, 2006; Carbon et al., 2003; Carbon et al., 2004; Palmer et al., 2009). However, extended motor practice alone, while being effective during the early phases of sensory motor learning, may not be sufficient during late phases that involve automatization (Karni et al., 1998a; Wu et al., 2015). Augmented feedback provided as knowledge of results, alone or in combination with knowledge of performance, also improved motor skill retention. Persons with PD experience proprioceptive (Adamovich et al., 2001; Konczak et al., 2009) and central processing 21 integration impairments (Contreras-Vidal and Gold, 2004; Schindlbeck and Eidelberg, 2018; Tahmasian et al., 2017) as well as attentional deficits (Watson and Leverenz, 2010). Augmented feedback could therefore help these patients activate their cognitive reserve to use a more volitional mode of action (Ferrazzoli et al., 2018) as well as to focus on the extrinsic information of the task during practice to enhance long-term skill retention (Chiviacowsky et al., 2010; Wulf, 2013; Wulf et al., 2009).

## 5. Limitations

It is important to emphasize that most studies of the review included participants in the early symptomatic stages of PD. More studies are thus needed to determine if the task-specific deficits in skill retention found in this study remain present or are augmented in later stages of the disease. Given the limited number of studies available, it was not possible to conduct additional analyses investigating interactions between augmented feedback and task nature. Future studies should determine which forms, frequency, and focus (internal vs. external) (Wulf, 2013) of augmented feedback is more effective to improve skill retention across different (simple vs. complex) motor tasks (Wulf and Shea, 2002).

Since only three studies manipulated medication status (Hadj-Bouziane et al., 2013; Hayes and Hunsaker, 2015; Lahlou et al., 2022), we could not investigate the effects of antiparkinsonian medications on motor skill retention. Our sensitivity analyses, however, suggest that medication status did not affect retention (**Suppl. 4**). The effects that antiparkinsonian medications can have on motor skill acquisition and retention are complex and conflicting results have been reported (Carbon and Eidelberg, 2006; Carbon et al., 2003; Cools et al., 2001; Ghilardi et al., 2007; Marinelli et al., 2017a; Ruitenberg et al., 2015; Vaillancourt et al., 2013; Zhuang et al., 2013). More investigations are needed to elucidate the effects of these medications on motor memory consolidation processes and whether their potential modulating effects are similar across different types of motor tasks at different stages of the disease.

## 6. Conclusion

Persons with PD who perform sensory motor skills and sequential movements daily need to constantly adapt pre-existing motor routines to cope with the motor dysfunctions arising during the disease (Abbruzzese et al., 2009; Doyon, 2008; Wolpert et al., 2011). Similarly, these individuals suffer from gait disorders and postural instability (Allen et al., 2013; Bloem et al., 2001; Boonstra et al., 2008; Canning et al., 2014; Stolze et al., 2005), as well as speech and voice disorders (Miller et al., 2006; Ramig et al., 2011; Ramig et al., 2008). The results of this review confirm that people with mild to moderate PD have deficits in skill retention affecting primarily SMTs and VATs. These results underline the importance of developing targeted interventions to enhance motor memory processes to support long-term skill retention in this clinical population. Extended motor practice and augmented feedback might be valuable means to reduce deficits in motor skill retention, but more evidence is needed to confirm if these strategies are effective to improve different motor tasks.

## Data Availability

All data produced in the present study are available upon reasonable request to the corresponding author.

## Acknowledgements

We would like to thank the authors of the papers included in the review who provided additional information: Dr. Bo Foreman, Dr. Dagmar Sternad, Dr. Lisa Maurer, Dr. Alfredo Berardelli, MSc Soraya Lahlou, Dr. Ya-Yun Alice Lee, Dr. Evelien Nackaerts, Dr. Lucio Marinelli, Dr. Ann Smiley-Oyen, Dr. David Wright, Dr. Julien Doyon, Dr. Nicolas Nicastro, Dr. Karen Van Ooteghem, Dr. Jason Whitfield, Dr. Carolee Winstein, and Dr. Leszek Kaczmarek. We would also like to thank Alina Andretzky for reviewing an early version of this manuscript.

## Authors’ agreement

All authors approve the final version of the manuscript, which is the authors’ original work, has not received prior publication and is not under consideration for publication elsewhere.

## Data Statement

The data that support the findings of this study are available from the corresponding author upon reasonable request.

