## Supplementary materials for "Motor Skill Retention Impairments in Parkinson’s Disease: A Systematic Review with Meta-analysis"

**Supplementary 1. Search terms.**

| **Motor learning key Terms** | **PD key terms** | **Control key terms** |
| --- | --- | --- |
| "Motor Learning" OR "Motor Sequence" OR "Motor Sequence Learning" OR "Motor Skill Learning" OR "Sensorimotor Learning" OR "Motor Control" OR "Motor Adaptation" OR "Locomotor Adaptation" OR "Visuomotor Adaptation" OR "Visuo-Motor Adaptation" OR "Motor Skill" OR "Motor Task" OR "Motor Activity" OR "Motor Function" OR "Postural Task" OR Implicit OR "Implicit Learning" OR "Implicit Motor Learning" OR Explicit OR "Explicit Learning" OR "Sequence Learning" OR "Sequential Learning" OR "Procedural Learning" OR "Serial Reaction Time" OR Skill OR Practice OR Encoding OR Learning OR "Learning Task" | "Parkinson's Disease" OR "Parkinson Disease" OR Parkinson OR PD | "Healthy Control" OR "Healthy Controls" OR "Control Group" OR "Subject Controls" OR "Control Subjects" OR "Normal Subjects" OR "Age-Matched" OR "Age-Matched Participants" OR "Older Adults" OR "Healthy Adults" OR "Healthy Older Adults" OR "Healthy Elderly" |

**Supplementary 2. The NIH Quality Assessment Tool for Observational Cohort and Cross-Sectional Studies.**

***The NIH questions***:

Q1. Was the research question or objective in this paper clearly stated?
Q2. Was the study population clearly specified and defined?
Q3. Was the participation rate of eligible persons at least 50%?
Q4. Were all the subjects selected or recruited from the same or similar populations (including the same time period)? Were inclusion and exclusion criteria for being in the study prespecified and applied uniformly to all participants?
Q5. Was a sample size justification, power description, or variance and effect estimates provided?
Q6. For the analyses in this paper, was(were) the exposure(s) of interest measured prior to the outcome(s) being measured?
Q7. Was the timeframe sufficient so that one could reasonably expect to see an association between exposure and outcome if it existed?
Q8. For exposures that can vary in amount or level, did the study examine different levels of the exposure as related to the outcome (e.g., categories of exposure, or exposure measured as a continuous variable)?
Q9. Were the exposure measures (independent variables) clearly defined, valid, reliable, and implemented consistently across all study participants?
Q10. Was the exposure(s) assessed more than once over time?
Q11. Were the outcome measures (dependent variables) clearly defined, valid, reliable, and implemented consistently across all study participants?
Q12. Were the outcome assessors blinded to the exposure status of participants?
Q13. Was loss to follow-up after baseline 20% or less?
Q14. Were key potential confounding variables measured and adjusted statistically for their impact on the relationship between exposure(s) and outcome(s)?

**Supplementary 3.** **Meta-regression analyses for motor skill retention after single practice.**

The results of meta-regression analyses investigating the influence of moderators such as methodological quality (i.e., risk of bias), disease duration (years), severity (Hoehn & Yahr) motor function (Unified Parkinson’s Disease Rating Scale -UPDRS- part III) on skill retention after single practice are presented in the table below. None of these moderators had a significant effect on skill retention.

| ***Model*** | ***Independent variable*** | ***k*** | ***df*** | ***Q_Model_ / F*** | **R^2^** | **ß** | ***p-value*** |
| --- | --- | --- | --- | --- | --- | --- | --- |
| ***1*** | ***Risk of Bias – study quality*** | 47 | 2 | 3.6250 | 0.06 |  | 0.1750 |
|  | a. ***“Fair”*** | 34 |  |  |  | -0.1026 | 0.9180 |
|  | b. ***“Good”*** | 5 |  |  |  | -0.1024 | 0.7020 |
|  | c. ***“Poor”*** | 8 |  |  |  | -0.3981 | 0.0670 |
| ***2*** | ***DD*** | 36 | 1 | 0.1134 | 0.00 | 0.0143 | 0.7460 |
| ***3*** | ***H&Y*** | 30 | 1 | 0.9273 | 0.00 | 0.2146 | 0.3710 |
| ***4*** | ***UPDRS*** | 27 | 1 | 0.4432 | 0.00 | -0.0083 | 0.5200 |
| ***5*** | ***DD * UPDRS*** | 23 | 3-19 | 0.4784 | 0.16 |  | 0.6980 |
| ***6*** | ***H&Y * UPDRS*** | 18 | 3-14 | 0.4710 | 0.31 |  | 0.6870 |
| ***7*** | ***Motor Skill Acquisition*** | 47 | 2 | 4.7548 | 0.18 |  | 0.1190 |
|  | a. ***No difference*** | 43 |  |  |  | -0.1834 | 0.2660 |
|  | b. ***Favour of Control*** | 3 |  |  |  | 0.3465 | 0.2030 |
|  | c. ***Favour of PD*** | 1 |  |  |  | -0.8772 | 0.1440 |
| **Footnotes:** **DD** = Disease duration; **H&Y** = Hoehn & Yahr score; **UPDRS** = Unified Parkinson’s Disease Rating Scale part III. | | | | | | | |

**Supplementary 4. Sensitivity analyses exploring the effect of medication.**

Sentivity analysis were conducted to explore the effect of medication status on skill retention. To this end, conditions (i.e., effect sizes) in which the PD patients were practising and/or tested while “off” medication(Hadj-Bouziane et al., 2013; Hayes and Hunsaker, 2015; Kawashima et al., 2018; Lahlou et al., 2022; Platz et al., 1998) were removed from the main meta-analyses. Removing these conditions did not change the main results of the meta-analyses, suggesting that medication status did not affect skill retention after either single or extended practice.

***Single practice***: SMD = -0.21; 95% CI = -0.38, -0.05; *p* = 0.0123; N = 41; I^2^ = 44.4%

***Extended practice***: SMD = -0.07; 95% CI = -0.27, 0.14; *p* = 0.5286; N = 16; I^2^ = 0%

**Supplementary 5. Motor skill transfer after single practice.**

**Methods**: Motor skill transfer, which is an approach to investigate motor skill retention from the perspective of generalization(Schmidt et al., 2018), was investigated by grouping studies according to the number of practice sessions performed during acquisition (single vs. extended practice). To investigate the transfer of acquired motor skills to untrained tasks, we calculated group mean differences using consecutive retention and transfer test scores (e.g., transfer score 24h – retention score 24h). This method was adopted to reduce the influence that memory reactivation may have on motor memory processes(Dudai, 2004; Tronson and Taylor, 2007). Analyses were performed following the same procedures implemented in the main meta-analyses. However, we did not perform meta-regression analyses due to the limited effect sizes available.

**Results**: Motor skill transfer was investigated in seven(Marinelli et al., 2017; Mochizuki-Kawai et al., 2010; Nackaerts et al., 2020; Platz et al., 1998; Sidaway et al., 2016; Smiley-Oyen et al., 2003; Van Ooteghem et al., 2017) and one(Smiley-Oyen et al., 2012) studies after a single session and extended motor practice, respectively. Funnel and forest plots pertaining to motor skill transfer are reported below.

**Motor skill transfer: Funnel plot.**

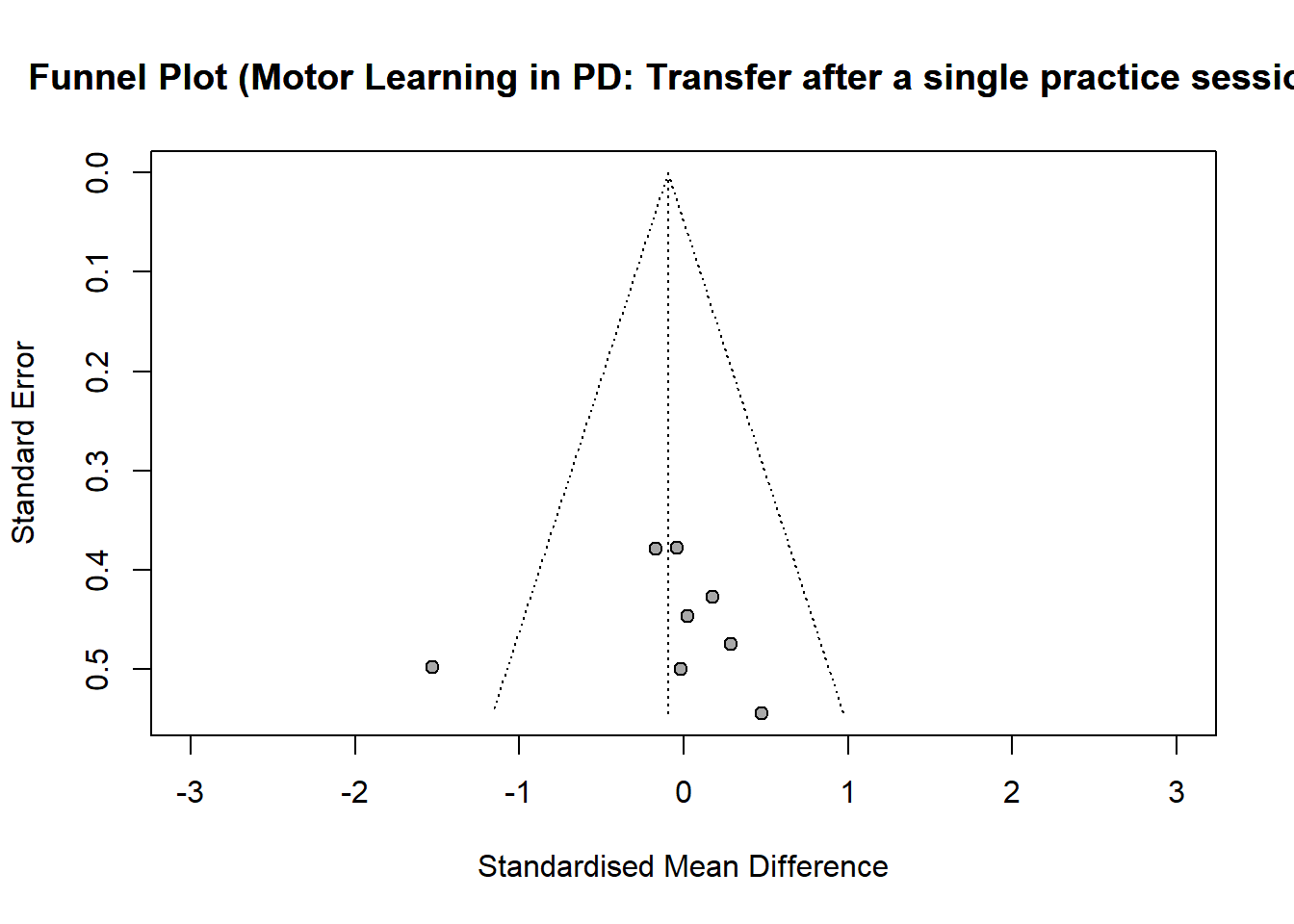

**Motor skill transfer: Forest plot.**

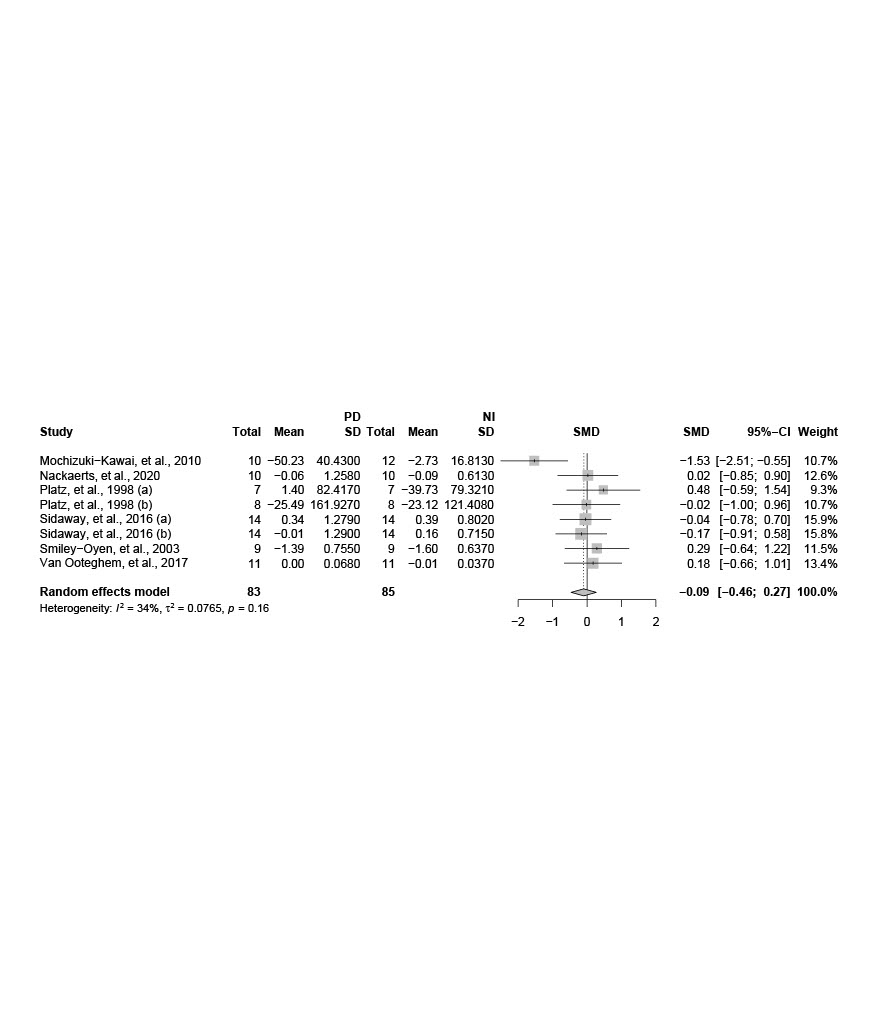

**Discussion**: Our results pertaining motor skill transfer following a single practice showed no difference between persons with the PD and NI group (SMD = -0.09; 95% CI = -0.46; 0.27; *p* = 0.6117; N = 8; I^2^ = 34.0%). These findings are relevant given that successful motor skill transfer would enable persons with PD to generalize and implement their motor skills in different contexts and tasks of daily living. However, our results should be considered preliminary and interpreted with caution for several reasons. First, only a limited number of effect sizes were available for the analysis (N = 8). Second, a few studies included in our review that observed motor skill transfer deficits in persons with PD(Isaias et al., 2011; Lee et al., 2016; Marinelli et al., 2017; Onla-Or and Winstein, 2008) could not be used in our analysis since it was not possible to obtain the data. For example, Isaias et al.,(Isaias et al., 2011) using a VAT, found deficits in motor skill transfer in persons with PD relative to NI individuals. Similarly, Onla-Or et al.,(Onla-Or and Winstein, 2008) observed that persons with PD have greater contextual interference than NI individuals in a SMT. Third, it is also plausible that our results were influenced by other factors such as the type of task implemented and the type of feedback provided. In addition to these considerations, previous studies have consistently observed that persons with PD have important deficits in performing acquired motor skills in new contexts and task-switching(Abbruzzese et al., 2009; Nieuwboer et al., 2009; Olson et al., 2019). Since motor skill transfer and generalization are important in PD, further research is needed to confirm and expand our preliminary results.

**Supplementary 6. Multiple effect sizes.**

It should be noted that 13 (Behrman et al., 2000; Dan et al., 2015; Harrington et al., 1990; Hayes and Hunsaker, 2015; Lahlou et al., 2022; Lee et al., 2019; Lin et al., 2007; Marinelli et al., 2009; Onla-Or and Winstein, 2008; Peterson et al., 2016; Platz et al., 1998; Sidaway et al., 2016; Smiley-Oyen et al., 2006) studies entered in the meta-analyses provided data to calculate multiple effect sizes, which were not pooled together to create a single comparison. This is because, among those studies, one used different motor tasks (Smiley-Oyen et al., 2006), three created different learning conditions by modifying either the complexity (Behrman et al., 2000; Harrington et al., 1990) or some parameters of the motor task (Peterson et al., 2016), and one study used different motor practice structures (blocked vs random) (Sidaway et al., 2016), which can alter motor learning processes (Carey et al., 2005; Chen et al., 2014; Lu et al., 2021; Schmidt et al., 2018). Furthermore, eight studies had more than two groups/experimental conditions, allocating different participants to each of them (Dan et al., 2015; Hayes and Hunsaker, 2015; Lahlou et al., 2022; Lee et al., 2019; Lin et al., 2007; Marinelli et al., 2009; Onla-Or and Winstein, 2008; Platz et al., 1998). Conversely, the retention scores of one study that used different active video games requiring similar motor and cognitive demands (Dantas et al., 2018) were pooled together to create a composite effect size.

**Supplementary 7. Flow diagram showing the flow of information through the different stages of the systematic review with meta-analysis.**

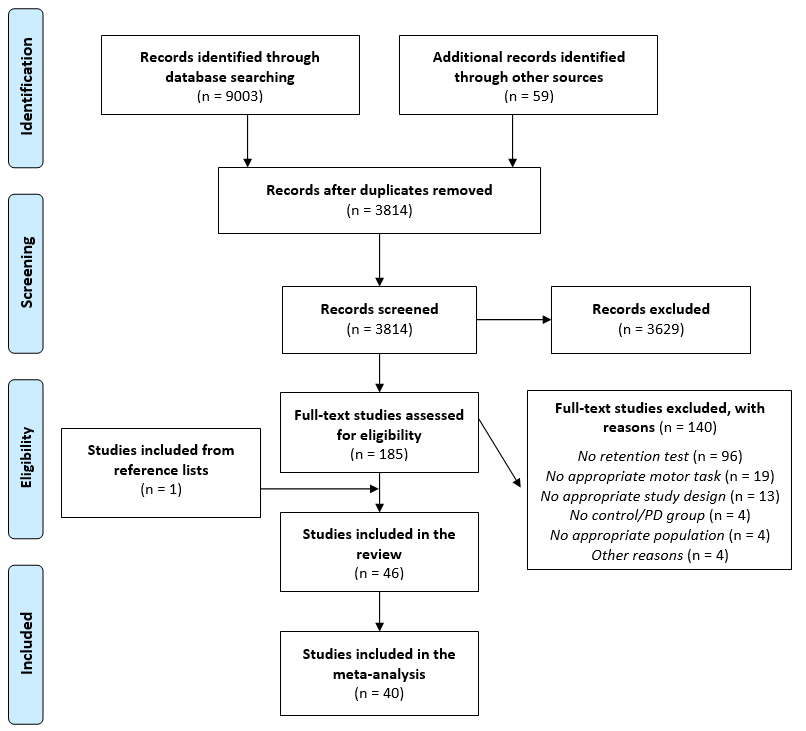

**Retention test**: Ninety-six studies were excluded because they did not have a retention test, or the latter was assessed less than one hour after the end of practice (eligibility criteria) (Abdel-Malek et al., 1988; Agostino et al., 2005; Agostino et al., 1992; Agostino et al., 1996; Alberts et al., 2000; Alberts et al., 1998; Ashoori et al., 2011; Barzgari et al., 2019; Bédard and Sanes, 2009; Beigi et al., 2016; Bondi and Kaszniak, 1991; Boonsinsukh et al., 2012; Broeder et al., 2019a; Caligiuri et al., 1992; Carbon and Eidelberg, 2006; Carbon et al., 2007; Carbon et al., 2003; Carbon et al., 2004; Carbon et al., 2010; Carey et al., 2002; Catala et al., 2016; Chen et al., 2016; Contreras-Vidal and Buch, 2003; Cressman et al., 2021; Cunnington et al., 1999; Deroost et al., 2006; Dominey et al., 1995; Dominey et al., 1997; Doyon et al., 1997; Fattapposta et al., 2000; Fernandez-Ruiz et al., 2003; Frith et al., 1986; Gamble et al., 2014; Geffe et al., 2016; Ghilardi et al., 2003; Ghilardi et al., 2007; Gobel et al., 2013; Guadagnoli et al., 2002; Heindel et al., 1989; Jackson et al., 1995a; Jackson et al., 1995b; Jordan and Sagar, 1994; Kelly et al., 2004; Kemeny et al., 2019; Kitahara et al., 2018; Krebs et al., 2001; Kwak et al., 2010, 2012; Linden et al., 1990; Lukos et al., 2010; MacAskill et al., 2002; Marinelli et al., 2010; Meissner et al., 2018, 2019; Meissner et al., 2016; Mentis et al., 2003a; Mentis et al., 2003b; Merritt et al., 2017; Messier et al., 2007; Mollion et al., 2011; Mongeon et al., 2013; Moreno Catalá et al., 2016; Nakamura et al., 2000, 2001; Nemanich and Earhart, 2015; Oates et al., 2013; Osman et al., 2008; Paquet et al., 2008; Pascual-Leone et al., 1993; Price and Shin, 2009; Rafal et al., 1987; Robertson and Flowers, 1990; Sarazin et al., 2002; Schendan et al., 2013; Smith et al., 2001; Smith and McDowall, 2006; Soliveri et al., 1992; Sprengelmeyer et al., 1995; Stefanova et al., 2000; Stephan et al., 2011; Thomas et al., 1996; Tremblay et al., 2010; Tzvi et al., 2021; van Tilborg and Hulstijn, 2010; Vandenbossche et al., 2013; Venkatakrishnan et al., 2011; Verschueren et al., 1997; Weiner et al., 1983; Werheid et al., 2003; Westwater et al., 1998; Wieczorek et al., 2011; Wilkinson et al., 2009; Wu and Hallett, 2005, 2008; Wu et al., 2015; Wu et al., 2010).

**No appropriate motor task**: Nineteen studies were excluded because they did not use an appropriate motor task/method to assess motor learning (Adam et al., 2011; Arroyo-Anll et al., 2015; Arroyo-Anllo et al., 2004; Beatty and Monson, 1990; Bellebaum et al., 2016; Chong et al., 2000; De Boer et al., 2016; DiFrancisco-Donoghue et al., 2015; Djamshidian et al., 2010; Filoteo et al., 2005; Grogan et al., 2017; Hodgson et al., 2013; Huang et al., 2017; Myers et al., 2003; Oishi et al., 2011; Oishi et al., 2010; Roncacci et al., 1996; Vakil and Herishanu-Naaman, 1998; Zokaei et al., 2020)

**No appropriate study design**: Thirteen studies used a study design that did not meet the inclusion criteria (Bello et al., 2008; Broeder et al., 2019b; Camacho et al., 2019; Chuma et al., 2006; Daum et al., 1996; dos Santos Mendes et al., 2012; Elangovan et al., 2018; Esculier et al., 2012; Flowers, 1976; Horiba et al., 2019; Lester et al., 2017; Moisello et al., 2015; Normand et al., 1993).

**No appropriate population**: Two studies used deep brain stimulation (Bédard and Sanes, 2011; Marcelino D.A. et al., 2019), one study included a control group with neurological conditions (Goldenberg et al., 1986), and another study had a control group with an age that differed significantly from the PD group (Ioffe et al., 2006).

**No control/PD group**: Four studies were excluded due to the lack of control/PD group (Adams et al., 2002; Chiviacowsky et al., 2012; Chung et al., 2020; Wu et al., 2004)

**Other reasons**: Four studies were excluded for other reasons (Bowen et al., 1973; Manuel et al., 2018; Sternad, 2015; Worringham and Stelmach, 1990) (e.g., duplicated data).

**Supplementary 8.** **NIH study quality assessment.**

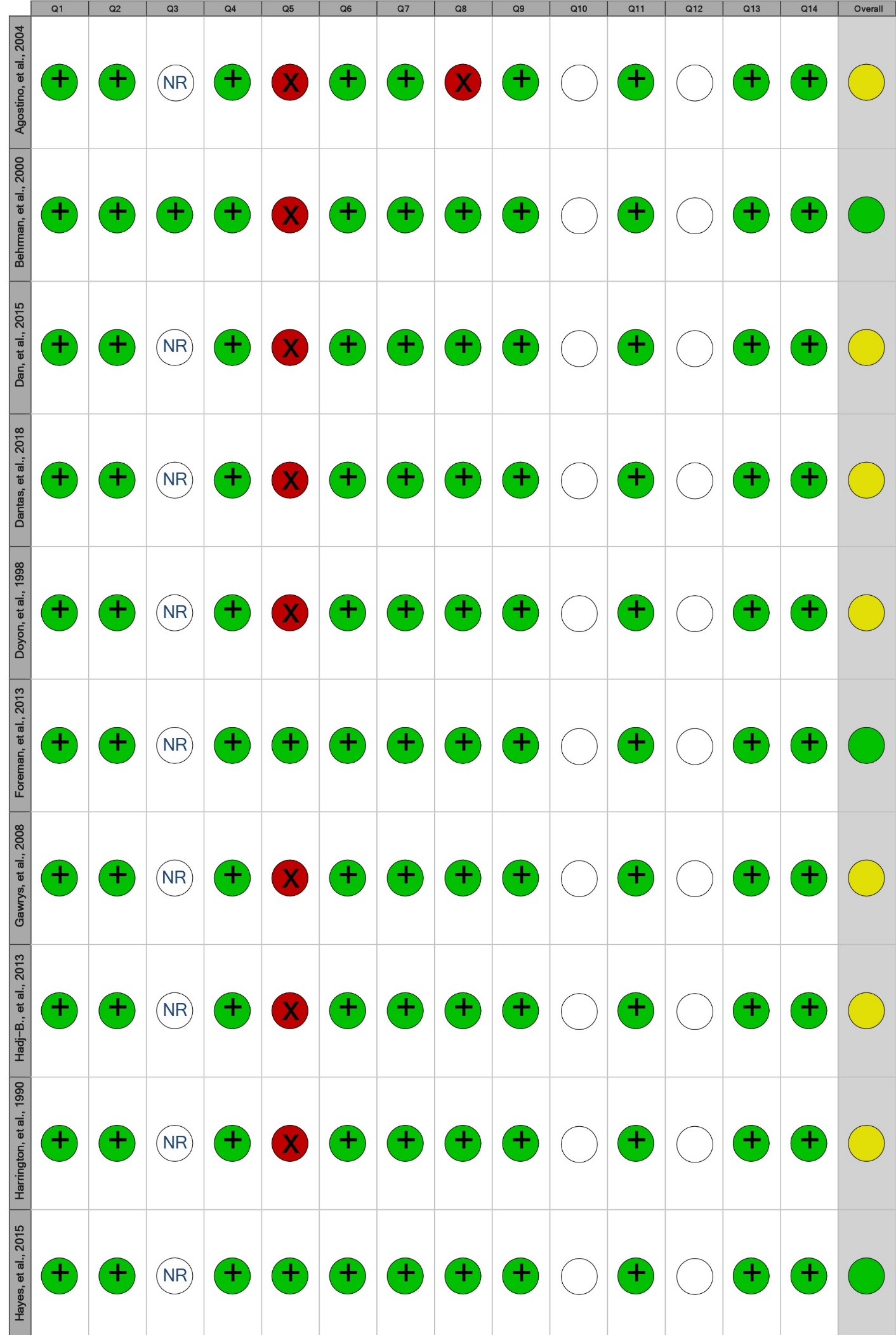

**Footnotes: • = No/Poor; • = Fair; • = Yes/Good; NR= not reported; " " = not applicable.**

**Supplementary 8.** *Continue*

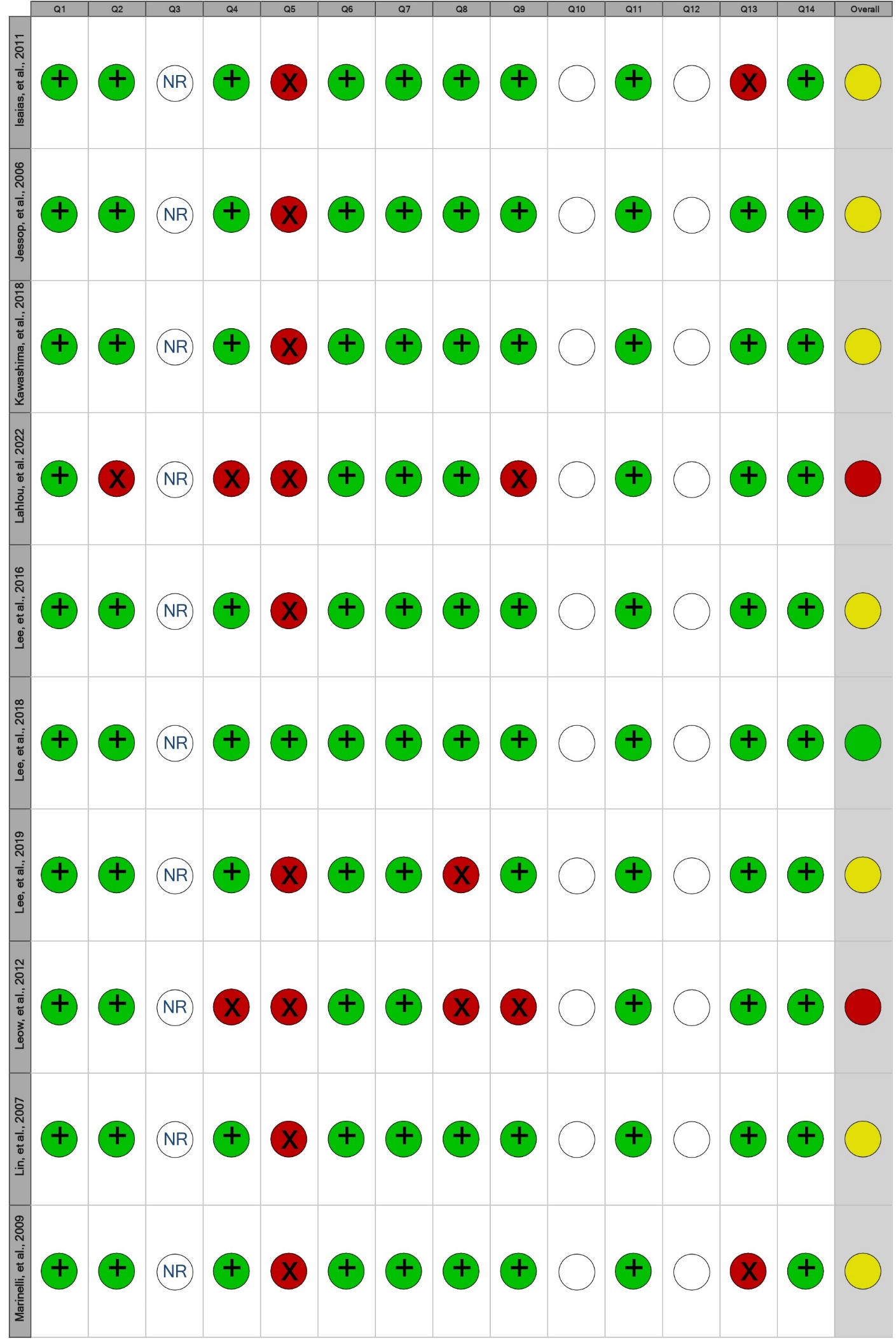

**Footnotes: • = No/Poor; • = Fair; • = Yes/Good; NR= not reported; " " = not applicable.**

**Supplementary 8.** *Continue*

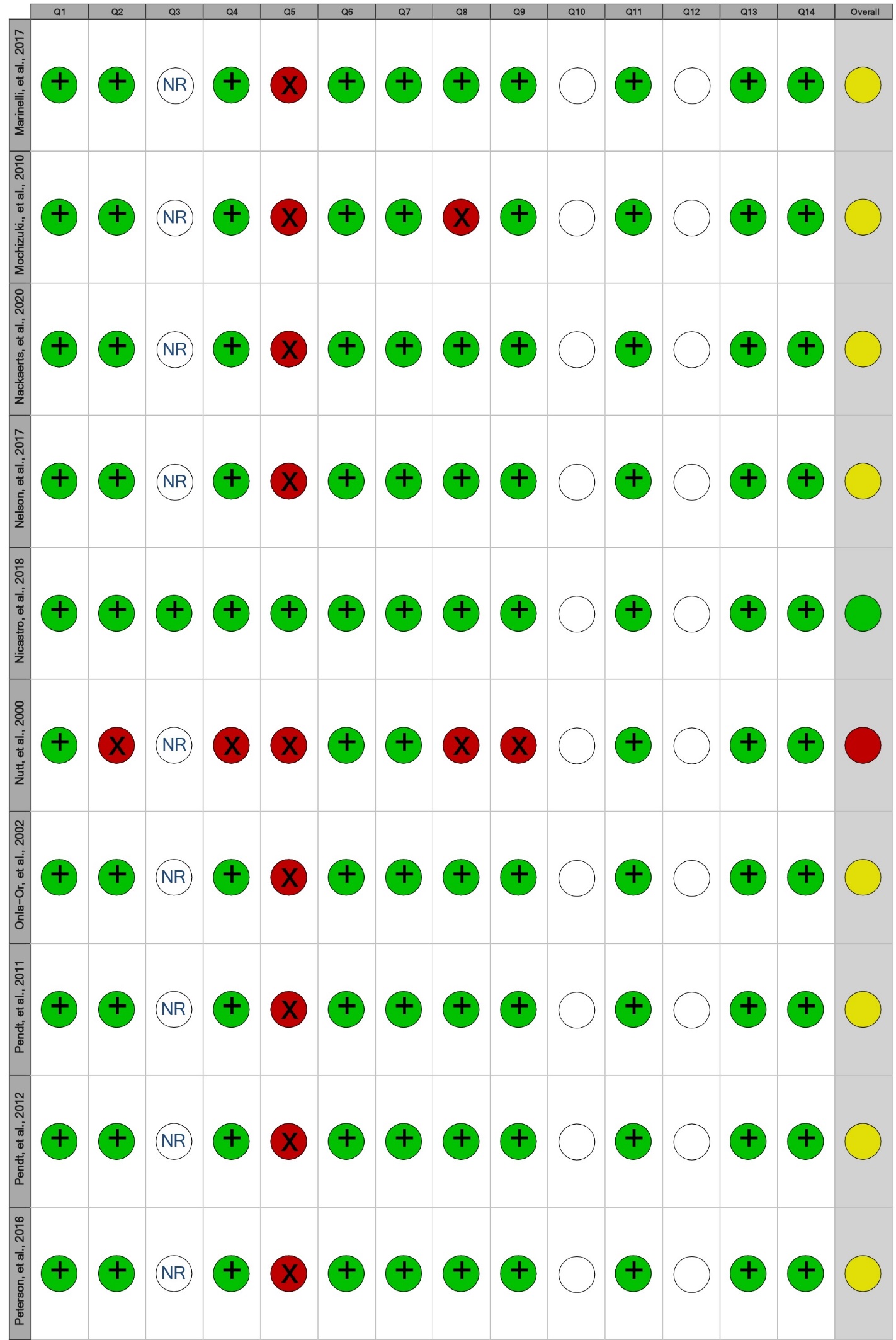

**Footnotes: • = No/Poor; • = Fair; • = Yes/Good; NR= not reported; " " = not applicable.**

**Supplementary 8.** *Continue*

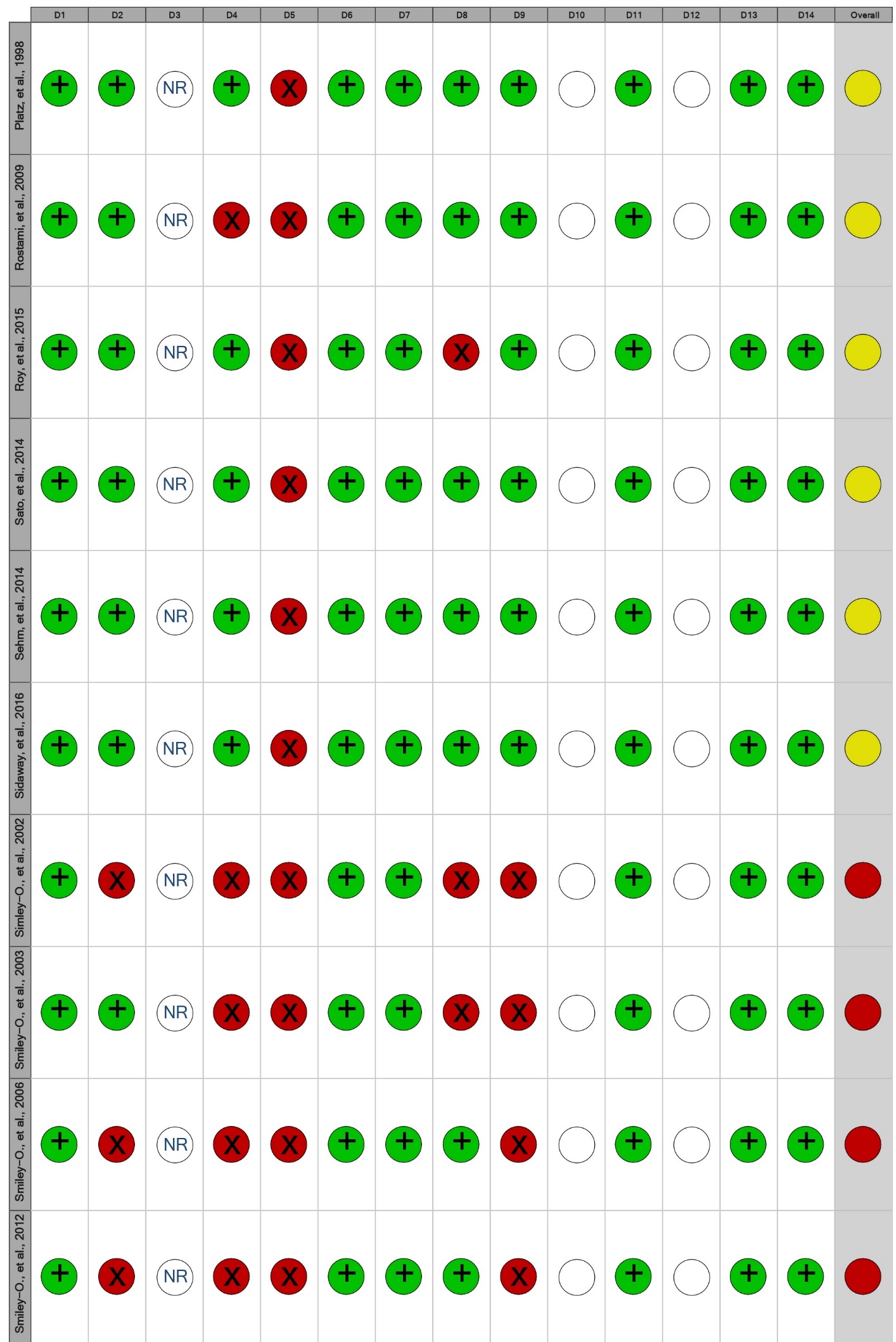

**Footnotes: • = No/Poor; • = Fair; • = Yes/Good; NR= not reported; " " = not applicable.**

**Supplementary 8.** *Continue*

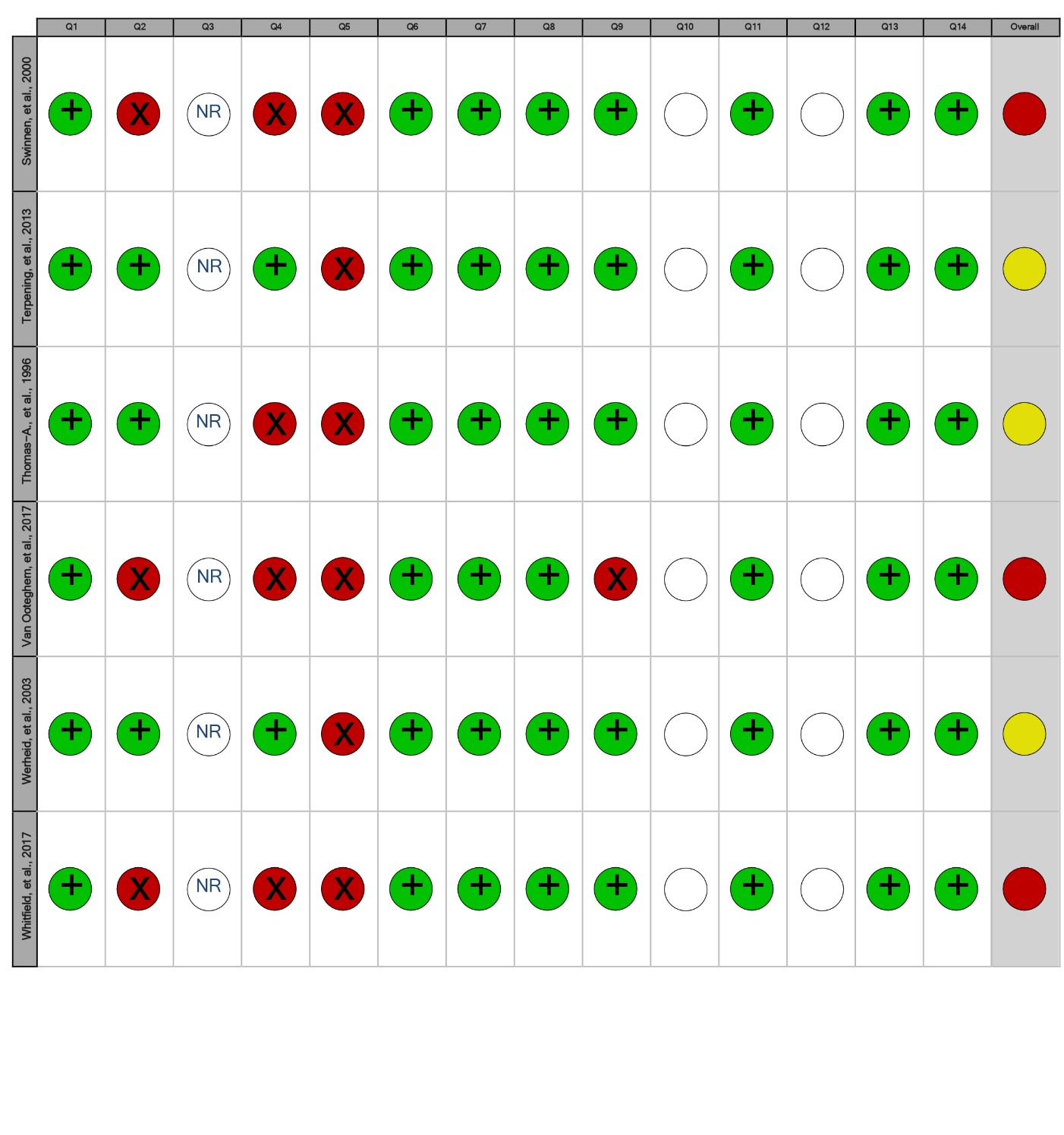

**Footnotes: • = No/Poor; • = Fair; • = Yes/Good; NR= not reported; " " = not applicable.**

**Supplementary 9.** **Funnels plots for motor skill retention after single and extended practice.**

Funnel plot – Motor skill retention after single practice.

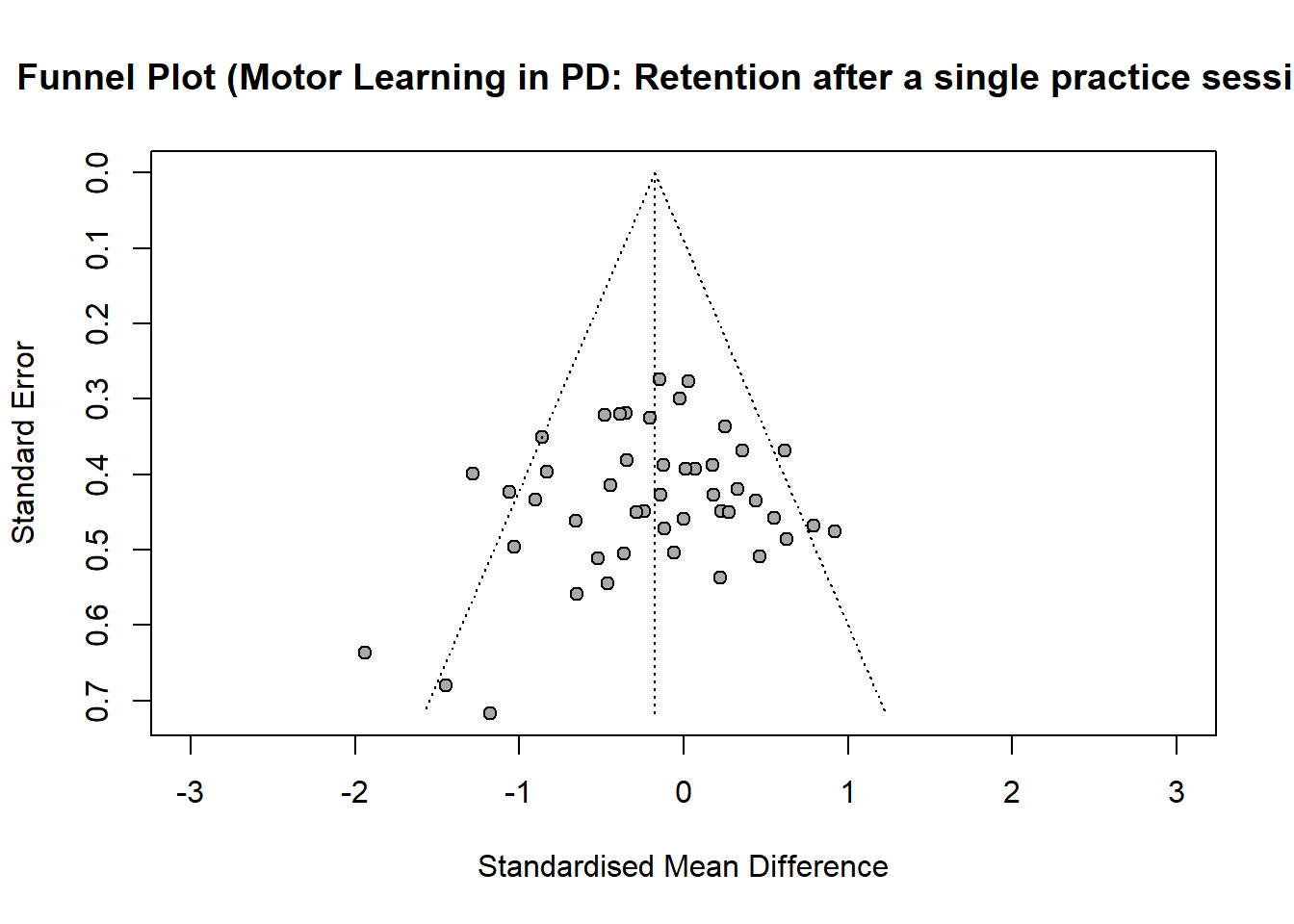

SMD

Funnel plots – Motor skill retention after extended practice.

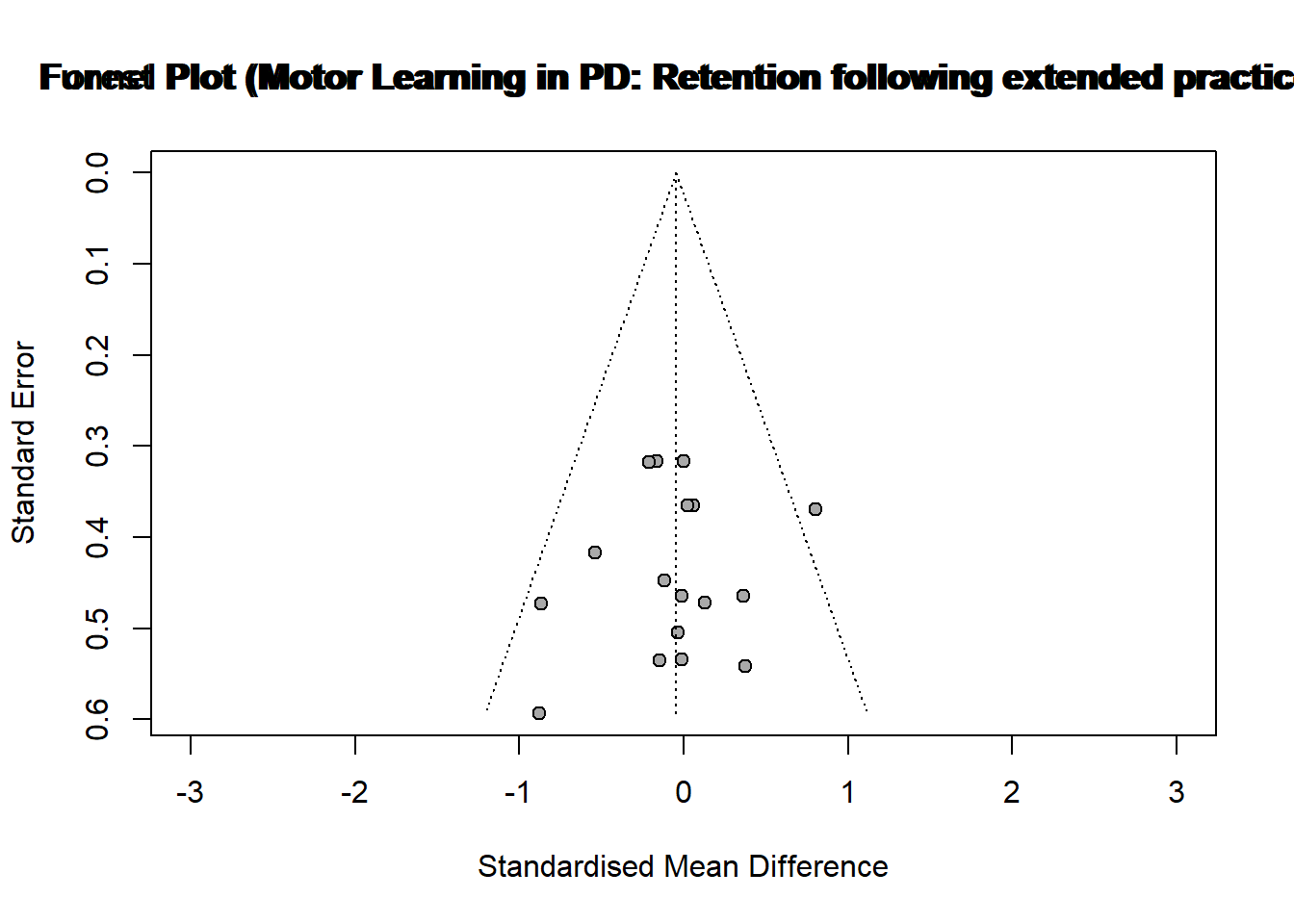

SMD

**Supplementary 10.** **Results of Egger’s tests.**

| **Egger’s test** | **Intercept** | **CI** | ***t-value*** | ***p*-*value*** |
| --- | --- | --- | --- | --- |
| *Motor skill retention after single practice* | -0.951 | -2.731; 0.829 | -1.046 | 0.301 |
| *Motor skill retention after extended practice* | -0.697 | -2.968; 1.574 | -0.602 | 0.556 |
